# Optimal control and cost-effectiveness analysis of mitigation strategies for monkeypox virus infection in the presence of routine interventions

**DOI:** 10.1101/2025.01.20.25320862

**Authors:** Samuel M. Naandam, Paul Chataa, Christopher Nikingbong, Agnes Adom-Konadu

## Abstract

Empirical evidence substantiates the pivotal role of vaccination in eradicating monkeypox virus (Mpox) infections. Mathematical modeling serves as a crucial tool for identifying strategies to curtail the disease’s proliferation. This study introduces a comprehensive mathematical framework for analyzing the dynamics of monkeypox transmission, incorporating imperfect vaccination and immunity degradation post-recovery. The theoretical constructs of positivity and boundedness are employed to confirm the model’s well-posedness. The next-generation matrix approach is utilized to calculate the control reproduction number (ℛ_*c*_), and the equilibrium points of the model are identified. The investigation demonstrates that the disease-free equilibrium is both locally and globally asymptotically stable, while the endemic equilibrium is proven to exhibit global asymptotic stability as well. Sensitivity analyses of parameters influencing ℛ_*c*_ are performed using Latin Hypercube Sampling (LHS) and Partial Rank Correlation Coefficients (PRCC). Furthermore, the model is extended to incorporate time-dependent interventions, including the administration of high-efficacy vaccines, the quarantine of exposed individuals, and the treatment of infected individuals. The model’s qualitative findings are reinforced through numerical simulations, which validate the effectiveness of various control strategies in suppressing monkeypox spread among susceptible, exposed, and infected populations. Simulations without vaccination controls reveal an initial surge in exposed and infected cases, followed by gradual containment over time. The findings highlight the critical importance of vaccination as a pharmaceutical intervention, though its effectiveness is hindered by challenges such as viral mutations, the diminishing efficacy of vaccines, and limited vaccination resources. These constraints underscore the necessity of adopting integrated intervention measures, especially as instances of reinfection post-vaccination and treatment of infected cases have been documented in several studies. Policymakers are thus encouraged to employ a balanced and pragmatic approach when easing other preventive measures for monkeypox. Additionally, a cost-effectiveness analysis was performed to determine the most economical strategy for controlling monkeypox transmission. The results indicate that the optimal control strategy involves combining high-efficacy vaccination with the quarantine of exposed individuals, demonstrating superior cost-effectiveness among the examined interventions.

## 1 Introduction

Monkey pox (Mpox) infection arises from the monkeypox virus (MPoxV), an enveloped, double-stranded DNA virus belonging to the Orthopoxvirus genus within the Poxviridae family. This virus comprises of two distinct genetic lineages, designated as clade I (previously known as the Central African clade) and clade II (formerly the West African clade). Clade I is associated with more severe illness and a significantly elevated case fatality rate, roughly 10%, compared to approximately 0.1% observed in infections caused by clade II. The worldwide emergence of monkeypox in May 2022 has underscored the considerable public health hazards posed by zoonotic diseases [1]. The virus predominantly transmits to humans via exposure to infected animals; however, human-to-human transmission may also transpire through multiple pathways [2, 3]. The disease unfolds in discrete phases, comprising a latency period, a prodromal phase, and an exanthematous stage distinguished by dermatological lesions [2, 4]. The persisting monkeypox outbreak has escalated into a public health crisis of international significance, with over 63, 000 cases documented, including its proliferation to regions such as the United Kingdom [5]. Although monkeypox cases are generally benign, high-risk groups including pediatric populations, gravid individuals, and those with immunosuppressive conditions may experience exacerbated disease severity [6, 7]. Historically, monkeypox outbreaks have predominantly afflicted marginalized communities in Africa, where the virus has become endemic [8, 9, 10]. Alarming concerns have arisen due to the attenuation of immunity following the discontinuation of smallpox vaccination, alongside the evolving epidemiological trajectory of the virus [11, 12]. To combat the monkeypox outbreak effectively, it is imperative to devise sophisticated disease modeling frameworks for strategic public health planning and response.

In prior years, limited scholarly focus has been directed toward monkeypox, resulting in a deficient comprehension of its transmission dynamics. Nevertheless, a select number of studies have attempted to employ mathematical modeling methodologies to elucidate the virus’s propagation mechanics. Mathematical modeling has been scrutinized by Al-Shomrani et al. [13] to elucidate the interplay between humans and animals in relation to the Monkeypox virus, while Okyere and Ackora-Prah [14] delve into the intricacies of human transmission mechanisms of the virus through a mathematical examination. The authors in Peter et al. [15] investigated the transmission dynamics of the monkeypox virus and determined that the isolation of infected individuals within the human population effectively mitigates disease propagation. Alharbi et al. [16] assessed the efficacy of therapeutic interventions and vaccination strategies as containment measures for monkeypox. In a study conducted by Michael et al. [17], the authors scrutinized a mathematical model for monkeypox that integrated surveillance as a pivotal control measure. Additional significant contributions are documented by El-Mesady et al. [18], Alshehri and Ullah [19], and Alzubaidi et al. [20]. The aforementioned mathematical models elucidate the influence of perfect vaccination and therapeutic interventions as control strategies for Mpox. However, it has been established that the Mpox vaccine, which is a vaccine for smallpox, does not confer complete protection to fully vaccinated individuals [21]. This means that individuals vaccinated with the Mpox vaccine can still get the infection if they come into contact with infected humans or rodents.

Decades of extensive investigation into smallpox therapeutics have culminated in the creation of treatments that may prove effective for mpox as well. For instance, an antiviral agent formulated for smallpox (tecovirimat) received approval from the European Medicines Agency in January 2022 for mpox treatment under extraordinary conditions [21]. Consequently, it is imperative to incorporate the concept of imperfect vaccination when modeling the transmission dynamics of Mpox infection. Thus the prevailing literature underscores the necessity for further investigation and exploration to attain a more profound understanding of the monkeypox phenomenon. This study aims to examine the transmission dynamics of monkeypox and its control within human populations by employing a classical deterministic model that incorporates imperfect vaccination and treatment as routine interventions.

The optimal control methodology serves as a potent instrument employed to ascertain the most effective and optimal control strategy for a system aimed at achieving a specified objective [22, 23, 24]. In the realm of optimal control theory, a system is conventionally transformed into a set of differential equations that delineate the system’s evolution over time, while the control strategy is represented as a function that maps the current state of the system to a control input. The primary aim of optimal control theory is to identify the control strategy that either minimizes or maximizes an objective function, all while adhering to constraints imposed on the system and the control input [25, 26]. Various methodologies exist for addressing optimal control problems, including both analytical techniques and numerical optimization strategies. Among the prevalent numerical methods utilized to resolve optimization challenges are Pontryagin’s maximum principle, dynamic programming, and model predictive control [27, 26]. The application of optimal control theory to establish optimal strategies for the eradication of infectious diseases, including Mpox, is documented in [28, 29].

In our investigation, we offer a novel contribution to the field by adopting a deterministic methodology that incorporates imperfect vaccination, thereby distinguishing our study from previous work by Addai et al. [30] and Li et al. [31], who employed the Caputo fractional derivative. Additionally, while Madubueze et al. [32] and Adepoju et al. [24] utilized a deterministic model and made commendable advancements, our research goes beyond their efforts by conducting more comprehensive analyses in optimal control and numerical simulations for the proposed strategies. Moreover, our study sets itself apart from the work of Alshehri et al. [19] and Adepoju et al. [24], who also used a deterministic model, by incorporating an imperfect vaccination strategy into our framework and performing an extensive cost-effectiveness analysis for the strategies employed. Our strong emphasis on these dimensions underscores the distinctive contributions and innovations that our study brings to the existing literature.

The subsequent divisions of this manuscript are articulated as follows: Section 2 expounds upon the derivation and conceptualization of the mathematical framework. Thereafter, Section 3 scrutinizes the foundational attributes of the model, whereas Section 4 is dedicated to a comprehensive disquisition of the model, encompassing the disease-free equilibrium, the control reproduction number, and the endemic equilibrium. The investigation into both local and global stability for the disease-free and endemic equilibria, alongside the sensitivity analysis of sthe model, is meticulously conducted in Section 5. Section 6 embarks on an in-depth exploration of an optimal control problem, elucidating its characterization, simulation, and analytical discourse. Numerical simulations pertinent to the optimal control problem are executed in Section 7, followed by an interpretative exposition of the model outcomes in Section 7.6. Conclusively, the manuscript culminates with a synthesis of the pivotal insights encapsulated in Section 8.

## 2 Basic Model Formulation

A deterministic, compartmentalized model delineating the transmission dynamics of the monkeypox virus, accounting for suboptimal vaccination efficacy and treatment limitations, is proposed. This model encompasses two distinct demographic groups: humans and rodents. The aggregate human population is divided into seven distinct compartments, comprising susceptible individuals *S*_*h*_(*t*), vaccinated individuals *V*_*h*_(*t*), exposed individuals *E*_*h*_(*t*), infected individuals *I*_*h*_(*t*), quarantined individuals *Q*_*h*_(*t*), individuals under treatment *T*_*h*_(*t*), and those who have recovered *R*_*h*_(*t*). The entire human population is denoted by *N*_*h*_(*t*), such that

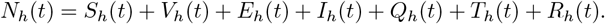

Similarly, the rodent population is divided into susceptible rodents *S*_*r*_(*t*), exposed rodents *E*_*r*_(*t*), and infected rodents *I*_*r*_(*t*), with the total rodent population represented as

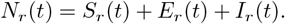

Susceptible humans are recruited into the population through birth and migration at an influx rate of Λ _*h*_. The probability of transmission from an infected rodent to a human is defined by the effective contact rate *χ*_1_, while *χ*_2_ represents the probability of a human contracting the virus after exposure to an infected human [26, 24]. Thus, the infection pressure exerted on the human population is given by

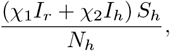

which consequently augments the exposed human compartment. The susceptible population is further expanded due to the attrition of immunity post-recovery at a rate *ζ*, erroneous diagnostic release of quarantined individuals at a rate *ϕ*_1_, and waning of vaccine-induced immunity at a rate *θ*_2_ [26, 24]. Additionally, a fraction of the susceptible population receives vaccination at a rate *θ*_1_. The immunization administered to at-risk individuals employs the smallpox vaccine, which demonstrates incomplete efficacy in conferring absolute protection against monkeypox infection [33, 34]. Accordingly, it is theorized that although vaccination diminishes predisposition to infection, it does not fully obliterate the risk [21, 33, 34]. This results in a modified force of infection for vaccinated individuals, represented by

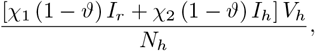

which further contributes to the exposed compartment. Here, *ϑ* ∈ (0, 1) signifies the vaccine efficacy, where *ϑ* = 0 indicates ineffectiveness, and *ϑ* = 1 denotes full 100% efficacy. The exposed category diminishes as individuals transition to either active or latent infections at rates *σ*_1_ and *σ*_2_, respectively [24]. The population of infected humans decreases due to a portion being treated at a rate *τ*_1_ [24], while spontaneous recovery occurs at a rate *τ*_2_ due to a robust immune response, and virus-induced mortality occurs at a rate *δ*_*h*_ [24]. We also assume that infected individuals who did not receive any form of treatment and did not recover naturally remain in the infected compartment. The rate at which quarantined individuals progress to an infectious state is *ϕ*_2_ [24]. Individuals undergoing treatment may either recover at a rate *τ*_3_ or experience treatment failure, resulting in reactivation of symptoms at a rate *ρ*. Individuals undergoing treatment for monkeypox are considered non-infectious once all scabs have fallen off and a fresh layer of skin has formed. This marks the end of the contagious phase of the disease. The virus is primarily spread through direct contact with lesions, body fluids, or contaminated materials like bedding and clothing during the active stage of the rash. As the lesions heal and new skin develops, the risk of transmitting the virus is effectively eliminated [35, 36]. All compartments within the human population are subject to natural mortality at a rate *µ*_*h*_.

For the rodent population, susceptible rodents enter the population at a rate Λ_*r*_. The probability of infection transmission from an infected rodent to another rodent occurs at an effective contact rate *χ*_3_, while exposed rodents progress to infection at a rate *σ*_3_ [26, 24]. Virus-induced mortality in rodents is indicated by *δ*_*r*_, and natural mortality by *µ*_*r*_. The general process of the model is described in Figure 1.

**Figure 1:**
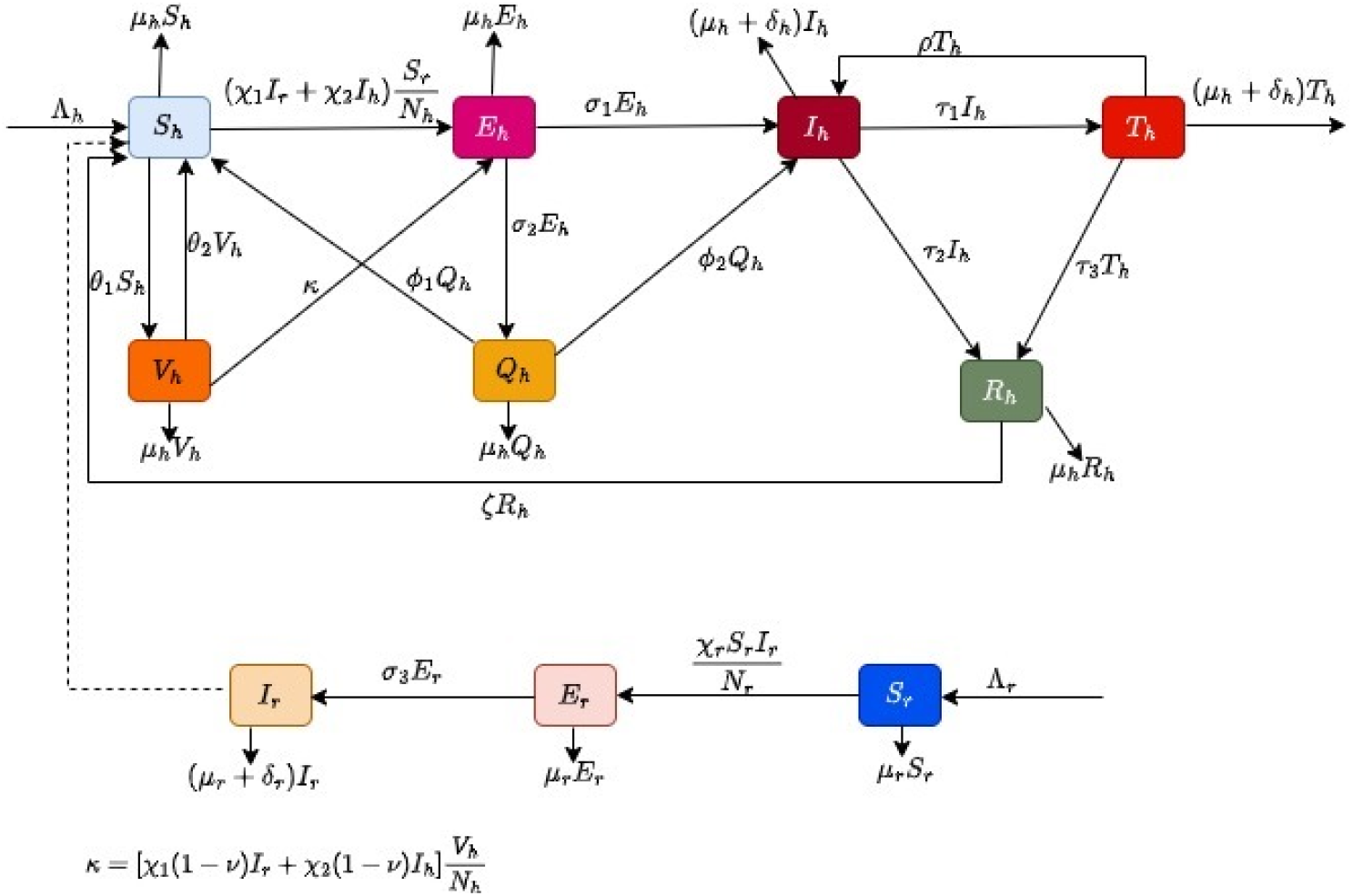
The outline illustrates the patterns of monkeypox spread

The mathematical formulation governing the transmission dynamics of monkeypox is thus articulated as fol-lows

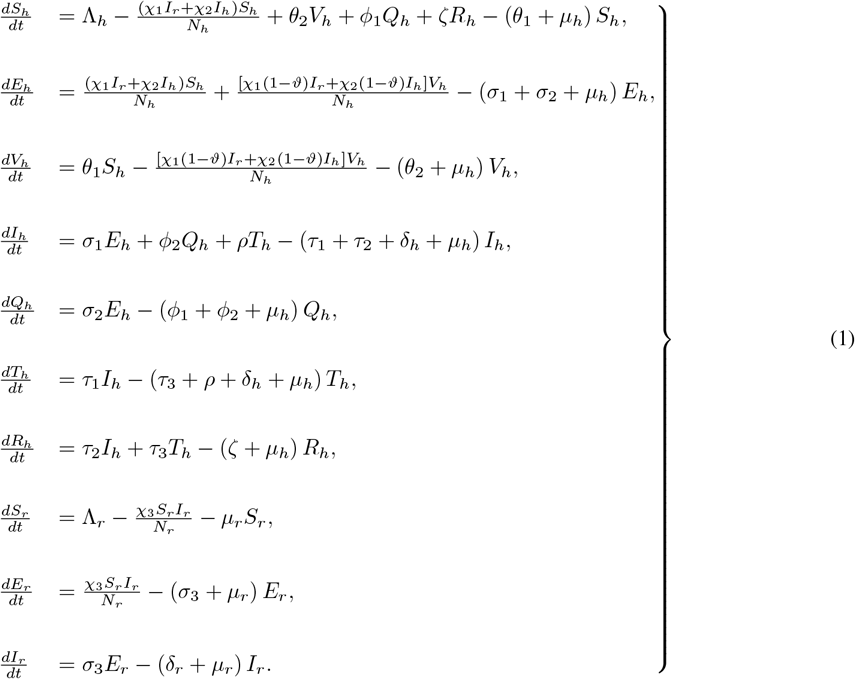

## 3 Basic model properties

In this section, we establish the positivity and boundedness properties of the solutions within the model framework.

### 3.1 Positivity of model solutions

For the system (1) to possess biological significance and maintain mathematical coherence, it is imperative to establish that all state variables within the model remain non-negative. Accordingly, under any positive initial conditions, the solutions of the model equations are guaranteed to remain positive throughout. Therefore, we present the following lemma to confirm the positivity of the solutions.

#### Lemma 1.

Assuming that the initial conditions and parameters of the system (1) are strictly positive, the solutions *S*_*h*_(*t*), *E*_*h*_(*t*), *V*_*h*_(*t*), *I*_*h*_(*t*), *Q*_*h*_(*t*), *T*_*h*_(*t*), *R*_*h*_(*t*), *S*_*r*_(*t*), *E*_*r*_(*t*), and *I*_*r*_(*t*) remain non-negative for all *t* ≥ 0.

*Proof*. Let us consider

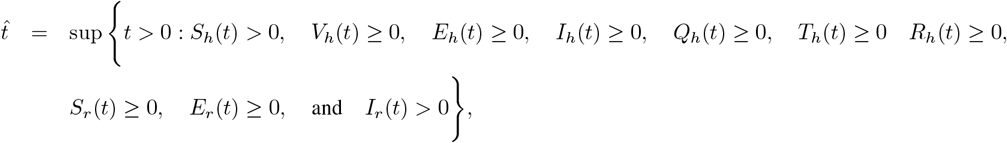

this implies that

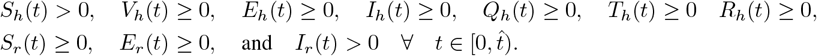

For the first equation in system (1), we have

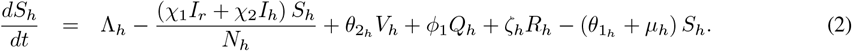

It follows from (2) that

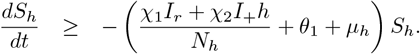

Hence,

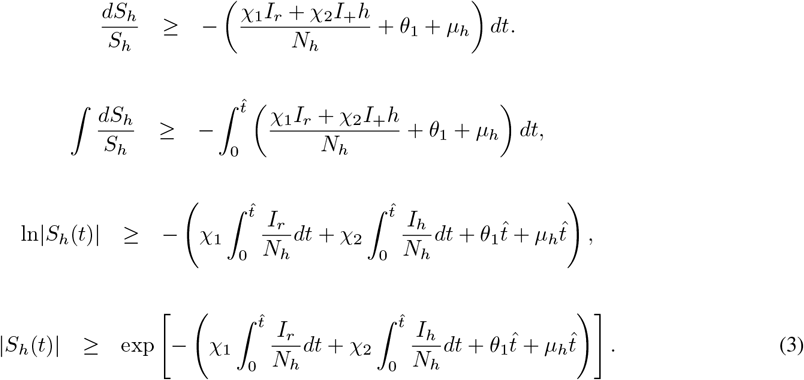

Since the exponential expression in the right side of (3) is always positive, it follows that

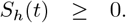

Also, for the third equation, we have

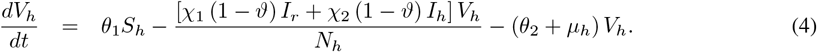

It follow from (4) that

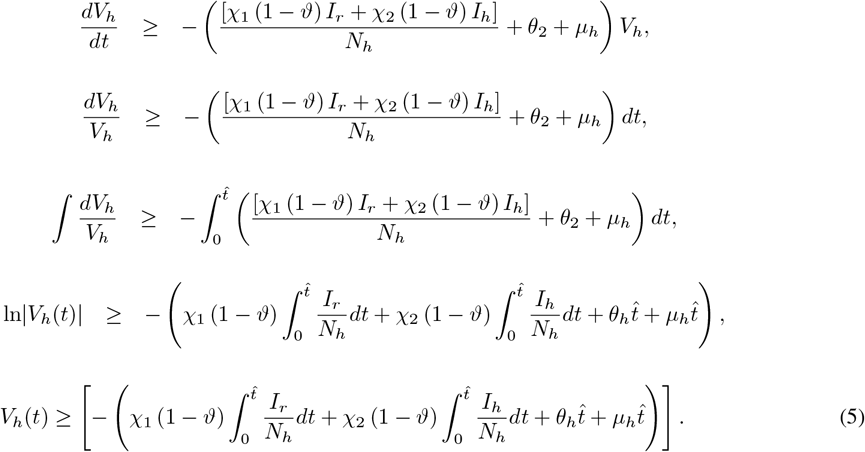

It follows from (5) that

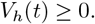

By employing an analogous methodology, the remaining equations yield

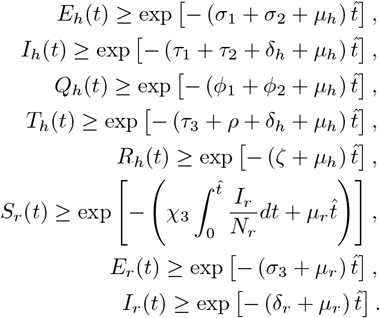

In conclusion, we have proved that the solutions *S*_*h*_(*t*) *>* 0 and *E*_*h*_(*t*) ≥ 0, *V*_*h*_(*t*) ≥ 0, *I*_*h*_(*t*) ≥ 0, *Q*_*h*_(*t*) ≥ 0, *T*_*h*_(*t*) ≥ 0, *R*_*h*_(*t*) ≥ 0, *S*_*r*_(*t*) ≥ 0, *E*_*r*_(*t*) ≥ 0, *I*_*r*_(*t*) ≥ 0 for all *t >* 0 given that *S*_*h*_(0) *>* 0 and *V*_*h*_(0) ≥ 0, *E*_*h*_(0) ≥ 0, *I*_*h*_(0) ≥ 0, *Q*_*h*_(0) ≥ 0, *T*_*h*_(0) ≥ 0, *R*_*h*_(0) ≥ 0, *S*_*r*_(0) ≥ 0, *E*_*r*_(0) ≥ 0, *I*_*r*_(0) ≥ 0.

### 3.2 Boundedness of model solutions

Under the premise that the initial parameters of the dynamical system (1) are defined as *S*_*h*_(0) *>* 0, *E*_*h*_ ≥ 0, *V*_*h*_ ≥ 0, *I*_*h*_(0) ≥ 0, *Q*_*h*_≥ 0, *T*_*h*_(0) ≥ 0, *R*_*h*_≥ 0, *S*_*r*_≥ 0, *E*_*r*_ ≥ 0, and *I*_*r*_ *≥* 0, and recognizing that the framework under analysis encapsulates the demographic dynamics of both human and rodent populations, it is sinferred that the aggregate human population, expressed as

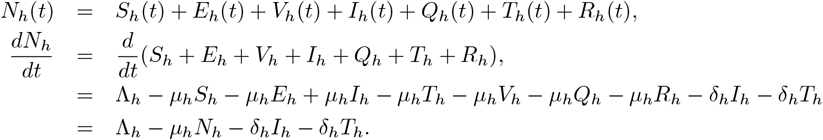

Hence, the rate at which the total human population is changing over time is given as

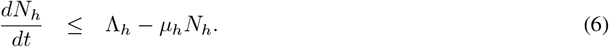

Therefore, the aggregate human population, *N*_*h*_(*t*), evolves in accordance with a steady influx rate (Λ_*h*_). Given that the system (1) tracks the dynamics of the human population, it is reasonable to postulate that all state variables and sparameters remain non-negative for all *t ≥* 0. Consequently, it can be demonstrated that all admissible solutions are uniformly bounded above by a certain threshold, beyond which neither human nor rodent population growth rates can escalate. The following conclusions are derived regarding the boundedness of the system (1).

#### Lemma 2.

The set defines the feasible region Ω_0_

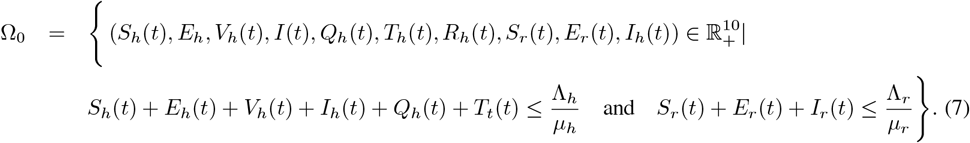

Since equation (6) is a standard first-order differential equation, we use the integration factor method to solve it. Thus,

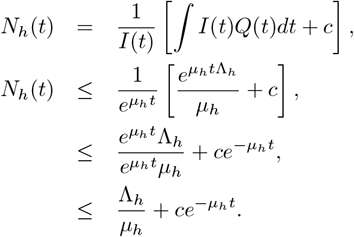

Where *Q*(*t*) = Λ_*h*_, *I*(*t*) = *e*^*µ*^ *h^t^* and as *t* → ∞ the solution for equation (6) is given as

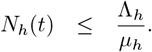

Hence, the human population is bounded above by 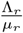. Similarly for the rodent population, we obtain

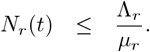

Therefore, the rodent population is bounded above by 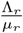. It follows that the region Ω_0_ is positively invariant and attracting with respect to system (1).

## 4 Model analysis

Within this section, we elucidate the disease-free and disease-persistence equilibrium states of the model framework, alongside the derivation of the control reproduction number.

### 4.1 Disease-free equilibrium and control reproduction number

The Disease-free Equilibrium (DFE) is a steady state of the system where the disease does not exist in the population. In this state all individuals in both human and rodent populations are either susceptible, vaccinated, or recovered. The infected and exposed compartments are empty (i.e *E*_*h*_ = 0, *I*_*h*_ = 0, *Q*_*h*_ = 0, *T*_*h*_ = 0, *E*_*r*_ = 0, *I*_*r*_ = 0). This equilibrium represents the baseline condition where the infection is absent, and the dynamics are governed only by demographic processes like recruitment, vaccination, and natural death. Thus, at the disease-free equilibrium, we have

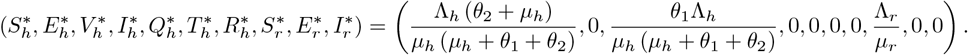

Subsequently, the Next Generation Matrix methodology, delineated by [37] and employed within the framework of [38, 39], is adopted to ascertain the control reproduction number of the constructed dynamical model. This metric encapsulates the mean quantity of subsequent Mpox cases incited by a solitary individual harboring the infection when introduced into a cohort of individuals wholly susceptible to the contagion. In other words, under the condition of an entirely uninfected population, the control reproduction number, symbolized as ℛ_*c*_, quantifies the aggregate of secondary infections engendered by a single Mpox-infected infected patient. The determination of ℛ_*c*_ necessitates a meticulous examination of the system of equations that delineate the genesis of novel infection alongside the transformations in their respective states. The provenance of these equations is comprehensively expounded by

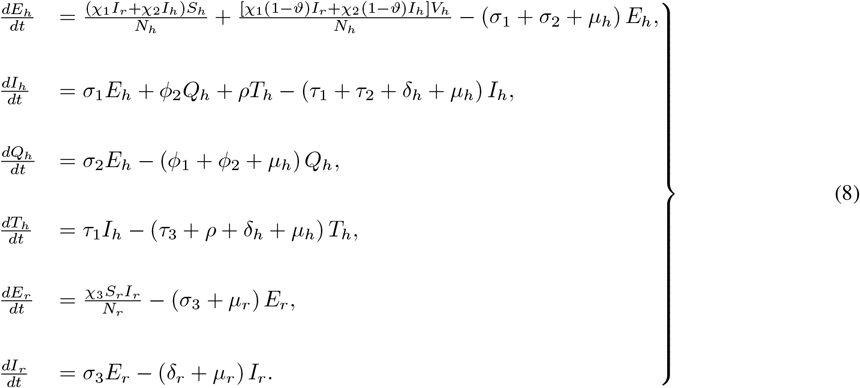

The assemblage of equations constituting system (8) is designated as the infected subsystem. According to established methodology, the initial step involves linearizing the infected subsystem around the disease-free equilibrium. By defining the vector

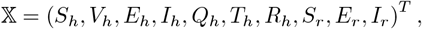

where *T* represents the transpose operation, the infected subsystem can subsequently be reformulated as

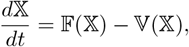

where 𝔽 𝕏 signifies the rate of emergence of new infections, and 𝕍 𝕏 captures transitions and removal processes within the infected states. It follows that

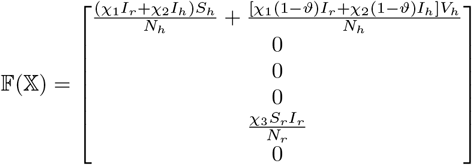

and

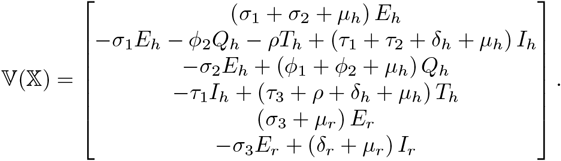

Taking the Jacobian of 𝔽 𝕏 at the disease-free equilibrium state, we have

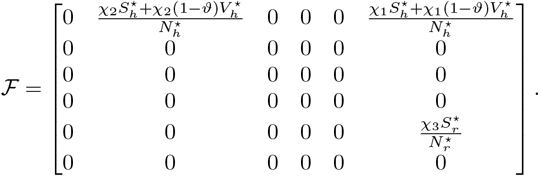

Also taking the Jacobian of 𝕍 𝕏 at the disease-free equilibrium, we obtain

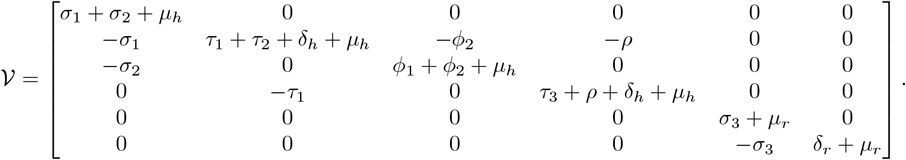

Thus,

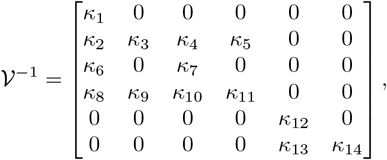

where

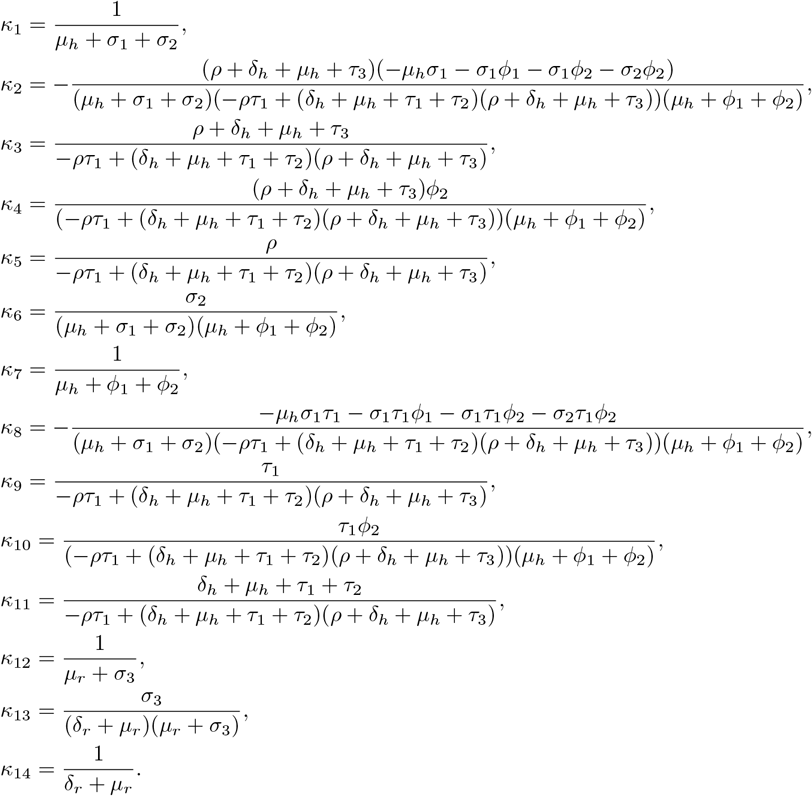

Hence *FV*^−1^ gives

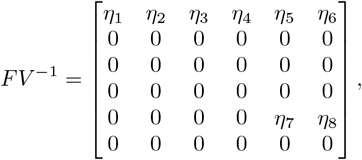

where

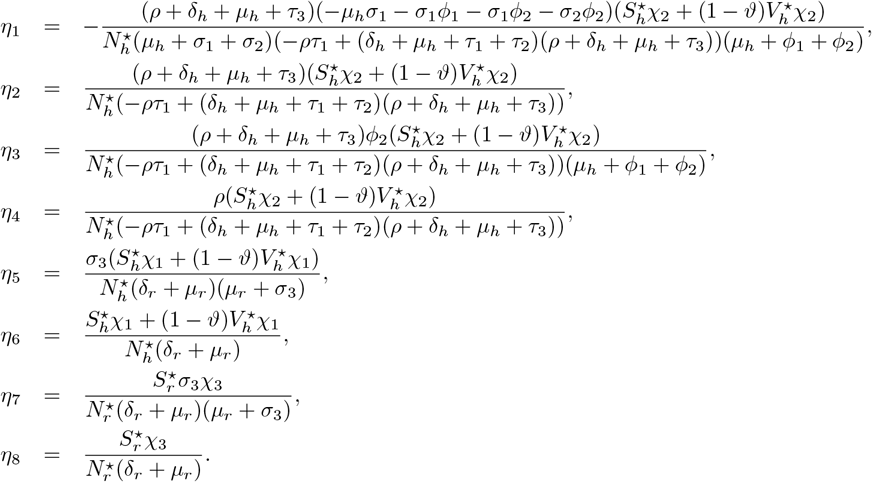

Therefore,

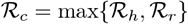

where

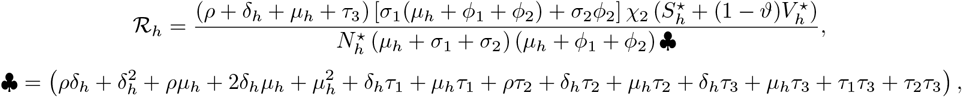

and

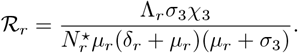

Consequently, given our focus on mitigating the propagation of the virus within the human populace, the study conducted herein evaluates all potential scenarios concerning the control reproduction number for the two populations, including instances where *ℛ*_*h*_ *> ℛ* _*r*_, *ℛ*_*h*_ *< ℛ*_*r*_, and *ℛ*_*h*_ = ℛ_*r*_. Throughout this investigation, the analysis has been undertaken with the primary aim of exploring strategies to curtail transmission to the human population under the condition where *ℛ*_*h*_ *<* 1 and *ℛ*_*r*_ *<* 1.

### 4.2 Disease-persistence equilibrium

In this section, we deduce the equilibrium points of disease persistence by simultaneously solving the governing mathematical framework (1) for the state variables 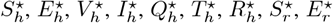 and 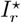. These equilibrium points represent steady-state configurations in which the Mpox infection endures within various demographic groups, signifying that eradication of the pathogen has not been achieved and the infection continues to exert influence across both human and rodent populations. For the Mpox infection to perpetuate within the host population, a non-zero viral reservoir is requisite. Consequently, the dynamics of the system are intricately dependent on the interplay of all state variables. At the equilibrium of disease persistence, previously unaffected humans and rodents acquire infection, thereby activating the respective infected compartments through interspecies interactions. Moreover, at the equilibrium of disease persistence, the infectious pressure exerted on susceptible populations are 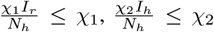 and 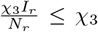 [39]. To rigorously scrutinize the theoretical ramifications of *ϑ* on the equilibrium distributions of infected individuals, we deliberate the scenario wherein 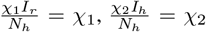 and 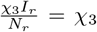, facilitating the derivation of explicit formulations for the endemic magnitudes of the state variables [39]. As such, the following equations must be fulfilled at this equilibrium

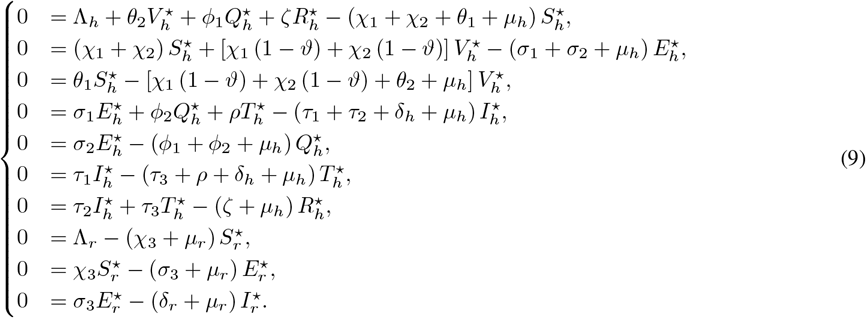

Employing the octaval, nonal, and decennial equations of (9), we ascertain

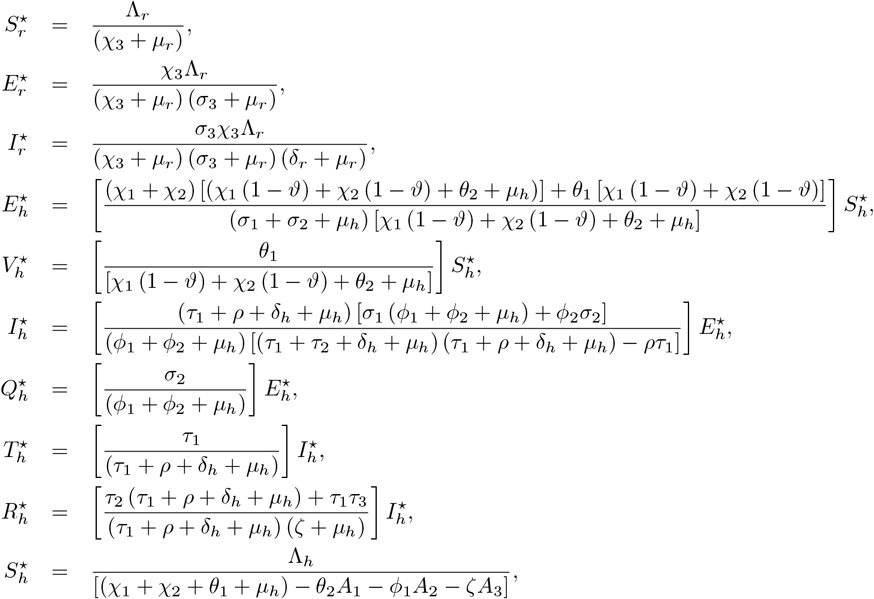

where

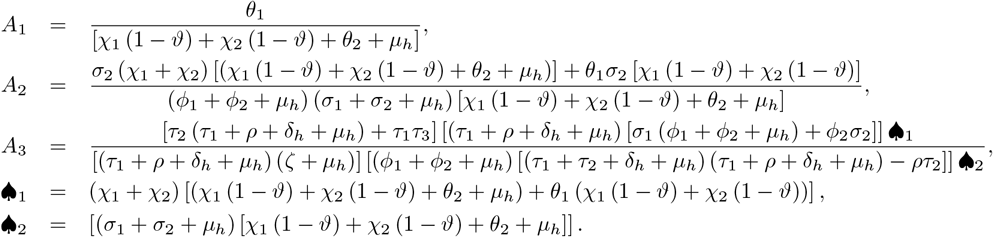

In this case, the solution exists and it is unique. From these conditions, we conclude that the endemic equilibrium solution is stable if and only if *ℛ* _*c*_ *>* 1 exhibits persistence of Mpox transmission in the population.

## 5 Stability Analysis

Within this section, we ascertain the proximal and overarching stability of the equilibrium devoid of infection.

### 5.1 Stability of the disease-free equilibrium

We ascertain the localized stability of the equilibrium devoid of disease by evaluating the eigenvalues of the linearized Jacobian matrix at the disease-free equilibrium.

#### Theorem 1.

The equilibrium free of infection for system (1) exhibits local asymptotic stability if *R*_*r*_ *<* 1 and Equation (10) holds true, and demonstrates instability if *ℛ* _*r*_ *>* 1 and Equation (10) does not hold.

*Proof*. We employ the Jacobian matrix corresponding to system (1) at the disease-free equilibrium and derive.

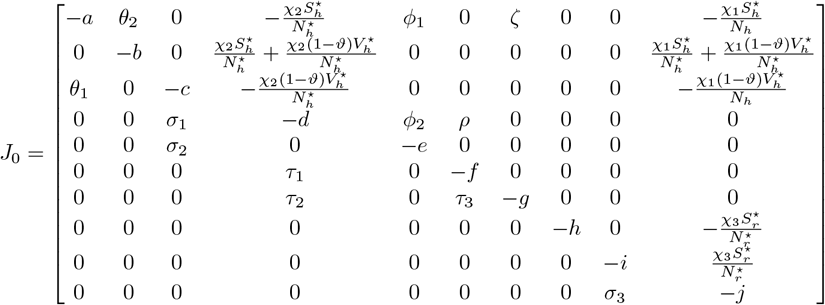

where

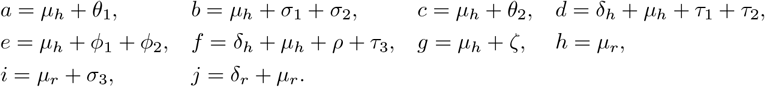

The eigenvaules from the jacobian matrix *J*_0_ are obtained to be

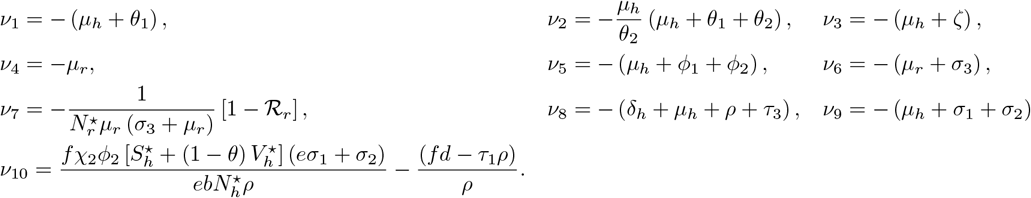

The eigenvalue *v*_10_ is negative if and only if

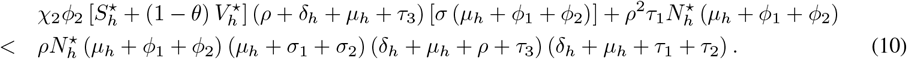

Since all the eigenvalues of the Jacobian matrix *J*_0_ have negative real parts when (10) is satisfied, it follows from the Routh-Hurwitz criterion that the disease-free equilibrium of system (1) is locally asymptotically stable.

### 5.2 Global stability of disease-free equilibrium

We further employ the methodology outlined in [40] and utilized in [38, 39] to establish the global stability of the disease-free equilibrium. This methodology is articulated in the theorem provided below.

#### Theorem 2.

If a model framework can be expressed as

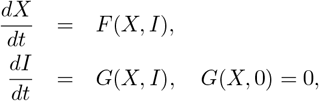

where *X* ∈ ℝ^*m*^ signifies the collection of uninfected compartments, and *I*∈ ℝ^*n*^ represents the infected compartments. Let *U*_0_ = (*X*^⋆^, 0) denote the disease-free equilibrium of the system. The following conditions, (*G*_1_) and (*G*_2_), must be satisfied to ensure global asymptotic stability.

(*G*_1_) For 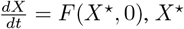 is globally asymptotically stable.

(*G*_2_) 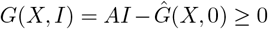 for (*X, I*) ∈ Ω, where *A* = *D*_1_*G*(*X*^⋆^, 0) is a Metzler matrix (its off-diagonal components are non-negative), and Ω represents the biologically meaningful and well-defined domain of the model.

Under these conditions, the fixed point *U*_0_ = (*X*^⋆^, 0) constitutes a globally asymptotically stable equilibrium of the Mpox infection model in system (1), provided that *ℛ*_*c*_ *<* 1.

#### Theorem 3.

The equilibrium devoid of disease for the model system,

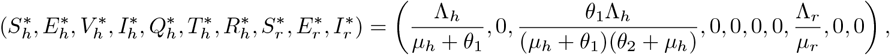

is globally asymptotically stable provided that ℝ_0_ *<* 1 and the conditions (*G*_1_) and (*G*_2_) are fulfilled.

*Proof*. From system (1), *X* ∈ ℝ^4^ = (*S*_*h*_, *V*_*h*_, *R*_*h*_, *S*_*r*_) and *I*∈ ℝ^6^ = (*E*_*h*_, *I*_*h*_, *Q*_*h*_, *T*_*h*_, *E*_*r*_, *I*_*r*_). Hence for condition (*G*_1_), we have

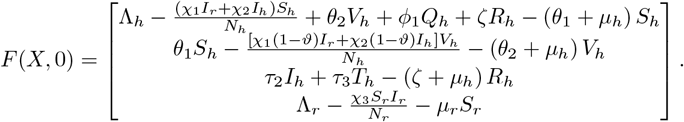

and

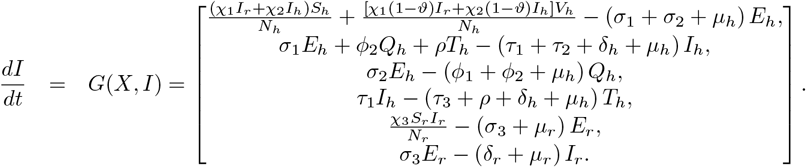

So, for the equilibrium *U*_0_ = (*X*^⋆^, 0), the system reduces to

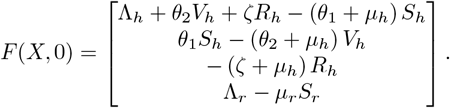

It follows that

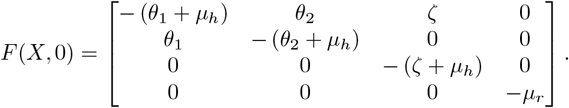

The eigenvaules from *F* (*X*, 0) matrix are obtained to be

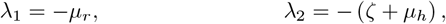

with a polynomial equation

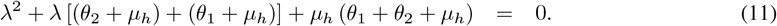

Given that all the coefficients of the characteristic polynomial in (11) are positive, the Routh-Hurwitz criterion ensures that the solutions to the characteristic polynomial possess negative real parts. Consequently, all eigenvalues are real and negative, implying that *X*^⋆^ remains globally asymptotically stable. Furthermore, utilizing Theorem 2 in the context of the Mpox model system (1) yields.

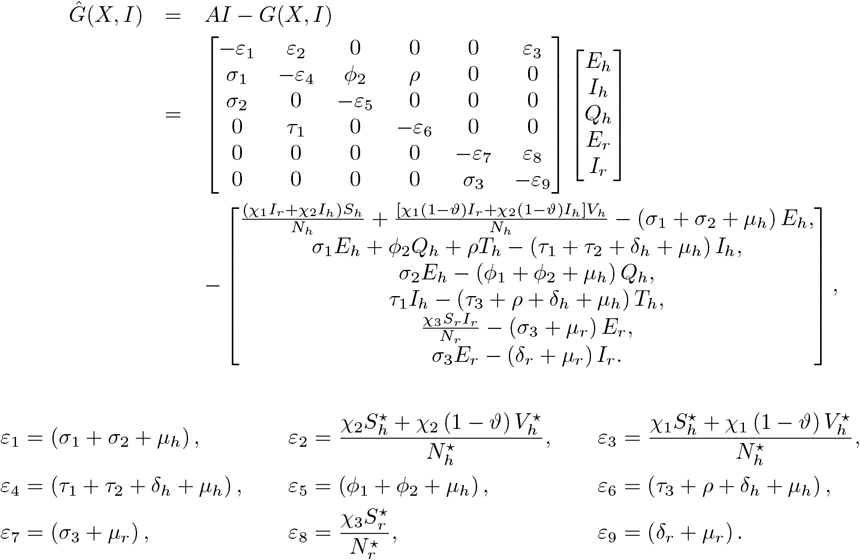

Hence, the context of Mpox model system (1) yields.

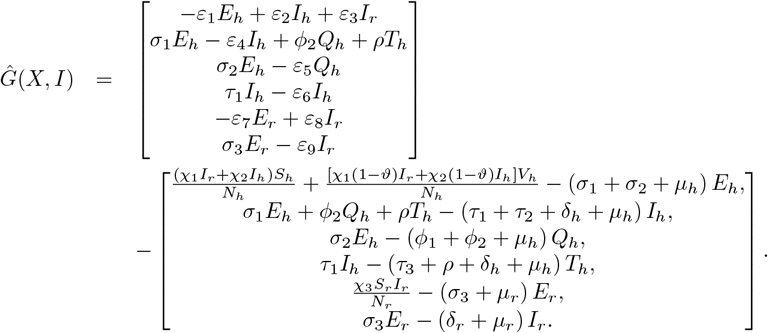

Therefore

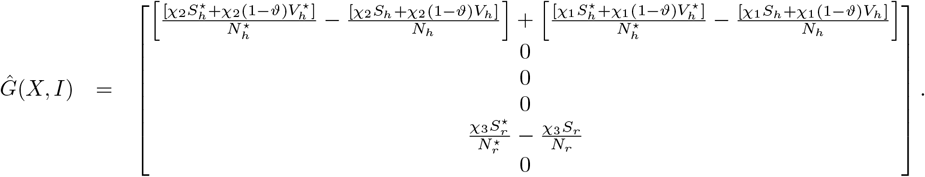

So, *A* is a Metzler matrix with non-negative off-diagonal elements. We observed that

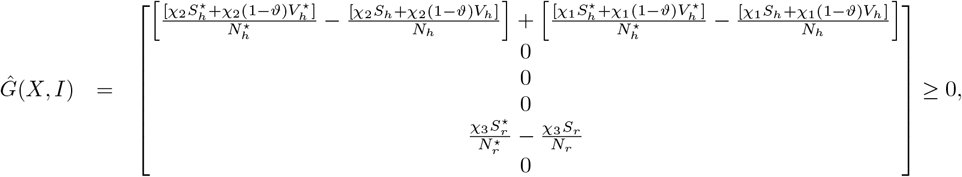

because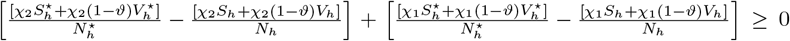 and 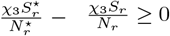 Thus, the disease-free equilibrium is globally asymptotically stable.

### 5.3 Global stability of endemic equilibrium

#### Theorem 4.

The inherent steady-state configuration of the Mpox dynamic system described in Equation (1) exhibits global asymptotic stability under the conditions *ℛ*_*h*_ *>* 1 (pertaining to human population) and ℛ _*r*_ *>* 1 (pertaining to rodent population). Conversely, it demonstrates instability when *ℛ*_*h*_ *<* 1 and ℛ_*r*_ *<* 1, as delineated in [34, 41].

*Proof*. In this study, we adopted a methodology analogous to that employed in [34, 42], utilizing a Logarithmic Lyapunov function formulated as

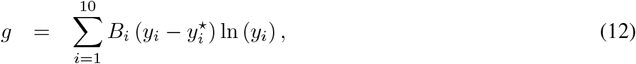

where *B*_*i*_ denotes a strictly positive constant, *y*_*i*_ represents the population magnitude of compartment *i*, and 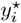 ssignifies the corresponding equilibrium value. It follows from Equation (12) that

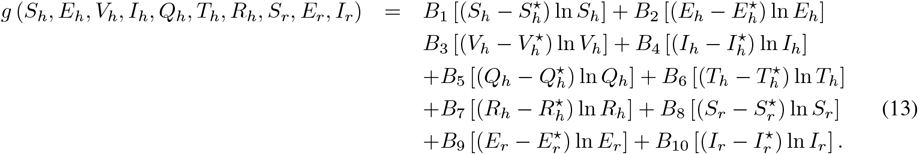

Upon taking the derivative of *g* with respect to time and simplifying we have,

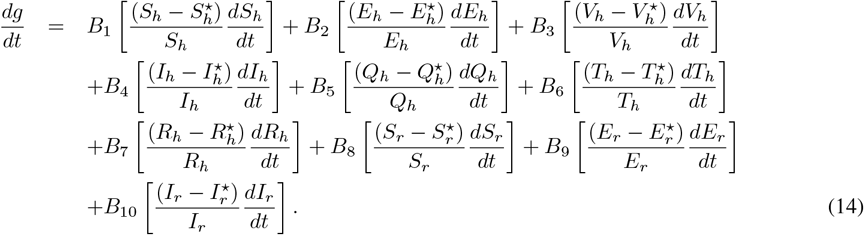

Substituting Equation (1) into Equation (14) we obtain

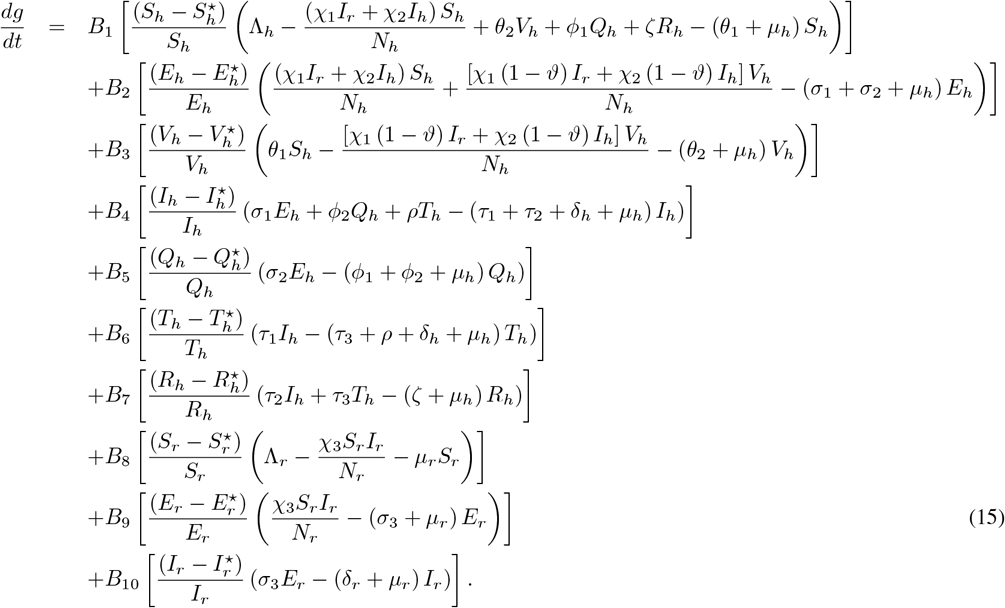

As demonstrated by Martcheva [43] and subsequently by Leandry and Mureithi [34], one of the conventional initial steps in this analysis involves substituting Λ _*h*_ and Λ _*r*_ with their corresponding expressions derived from the equilibrium equations, that is,

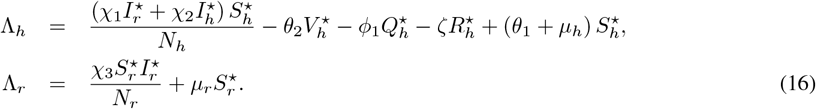

By substituting Equation (16) into Equation (15) and performing further algebraic simplifications, we obtain

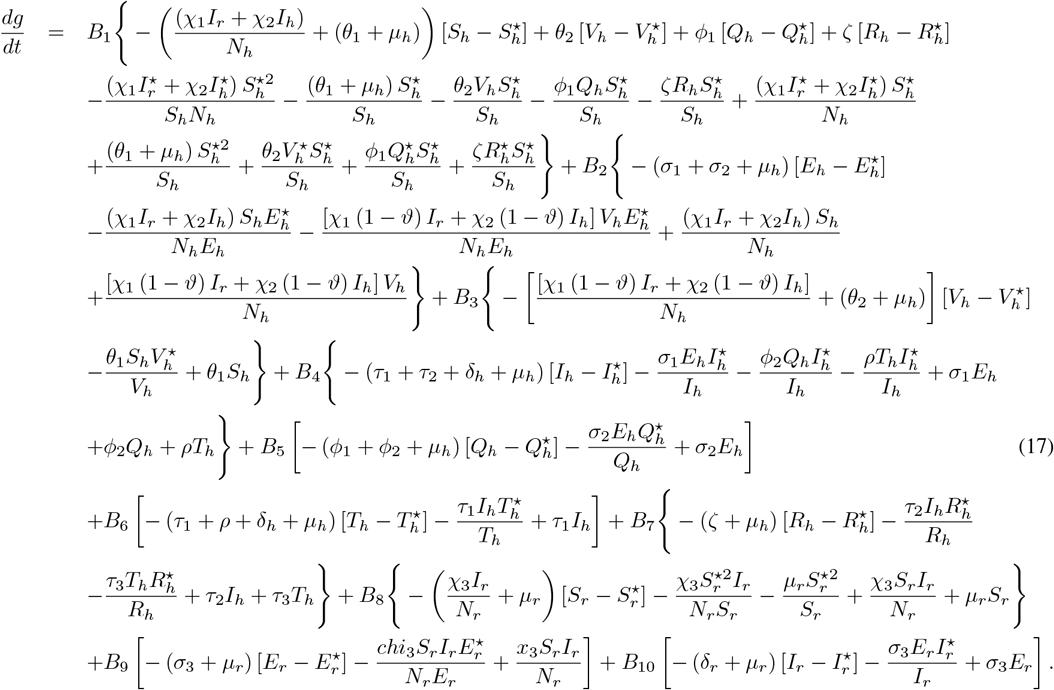

From Equation (17), we obtain

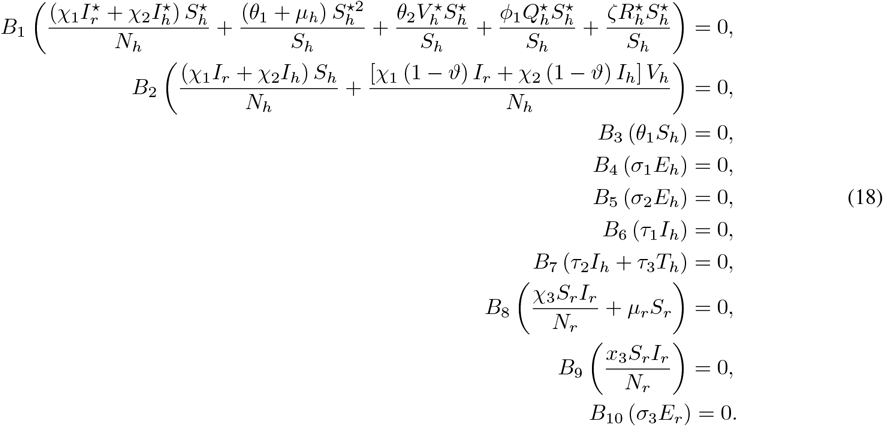

These expression in (18), when combined with additional terms in the product, results in negative contributions. We obtain the constants *B*_1_, *…, B*_10_ to be as follows.

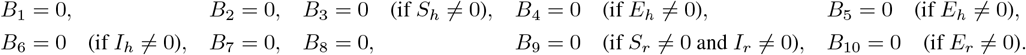

Subsequently, the Krasovskii–LaSalle invariance principle is employed. We focus on the subset where the Lyapunov function evaluates to zero, that is

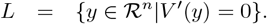

It is evident from Equation (15) that *V* ^*′*^ = 0 if and only 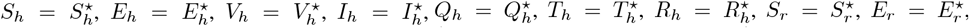 and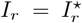. All other terms are negative when (*E*_*h*_, *V*_*h*_, *I*_*h*_, *Q*_*h*_, *T*_*h*_, *R*_*h*_, *S*_*r*_, *E*_*r*_, *I*_*r*_) *>* 0. Consequently, the endemic equilibrium is globally asymptotically stable under the conditions *R*_*h*_ *>* 1 and *R*_*r*_ *>* 1. This outcome substantiates and validates the theorem.

### 5.4 Sensitivity and uncertainty analysis

The sensitivity analysis elucidates the extent to which the parameters of the model exert influence over the controlled reproduction number *ℛ*_*c*_. A sensitivity assessment of *ℛ*_*c*_ concerning the model parameters was conducted utilizing the methodology articulated in [42]. The normalized forward sensitivity index of *ℛ*_*c*_ is contingent upon the differentiability of *ℛ*_*c*_ with respect to a specific parameter, denoted *k*, and is mathematically expressed as:

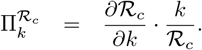

Subsequently, the methodologies of Latin Hypercube Sampling (LHS) and Partial Rank Correlation Coefficients (PRCC) are utilized to pinpoint the parameters within the model that wield the most substantial influence, with *R*_*c*_ functioning as the dependent variable. The primary aim of this inquiry is to elucidate the extent to which various parameters shape the model’s outputs. Parameters exhibiting high sensitivity necessitate more precise estimation, as minor perturbations can precipitate significant alterations in outcomes [44, 45, 46]. Conversely, parameters with low sensitivity demand less stringent estimation, given that small fluctuations in these parameters yield negligible impacts on the results [45]. Parameters attaining PRCC values exceeding 0.50 are classified as highly positively correlated with the dependent variable, whereas those with values below− 0.50 are designated as highly negatively correlated [44, 45, 46]. The PRCC analysis encompasses parameters such as *ρ, δ*_*h*_, *µ*_*h*_, *τ*_3_, *σ*_1_, *σ*_2_, *ϕ*_1_, *ϕ*_2_, *χ*_2_, *τ*_1_, *τ*_2_, Λ_*h*_, *θ*_1_, *θ*_2_ for *ℛ*_*h*_ and Λ_*r*_, *σ*_3_, *χ*_3_, *µ*_*r*_, *δ*_*r*_ for ℛ_*r*_. Although the analysis was conducted over five distinct intervals, the parameters demonstrated consistent effects on the dependent variable across all time frames. Consequently, a single representative plot is presented, as illustrated in Figures 2 and 3. The results reveal that the eight parameters exerting the most pronounced influence on the response function (ℛ _*c*_) are *δ*_*h*_, *µ*_*h*_, *χ*_2_, Λ_*h*_, Λ_*r*_, *χ*_3_, *µ*_*r*_, *δ*_*r*_. Based on the PRCC values, parameters *ρ, σ*_1_, *ϕ*_2_, *χ*_2_, Λ_*h*_, *θ*_2_, Λ_*r*_, *σ*_3_, *χ*_3_ exhibit a positive relationship with (ℛ_*c*_), implying that an augmentation (or diminution) in these parameters results in a corresponding increase (or decrease) in (*ℛ*_*c*_). In contrast, parameters *δ*_*h*_, *µ*_*h*_, *τ*_3_, *σ*_2_, *ϕ*_1_, *τ*_1_, *τ*_2_, *θ*_1_, *µ*_*r*_, *δ*_*r*_ display a negative correlation with (ℛ_*c*_), indicating that an elevation in these parameters diminishes (ℛ_*c*_).

**Figure 2:**
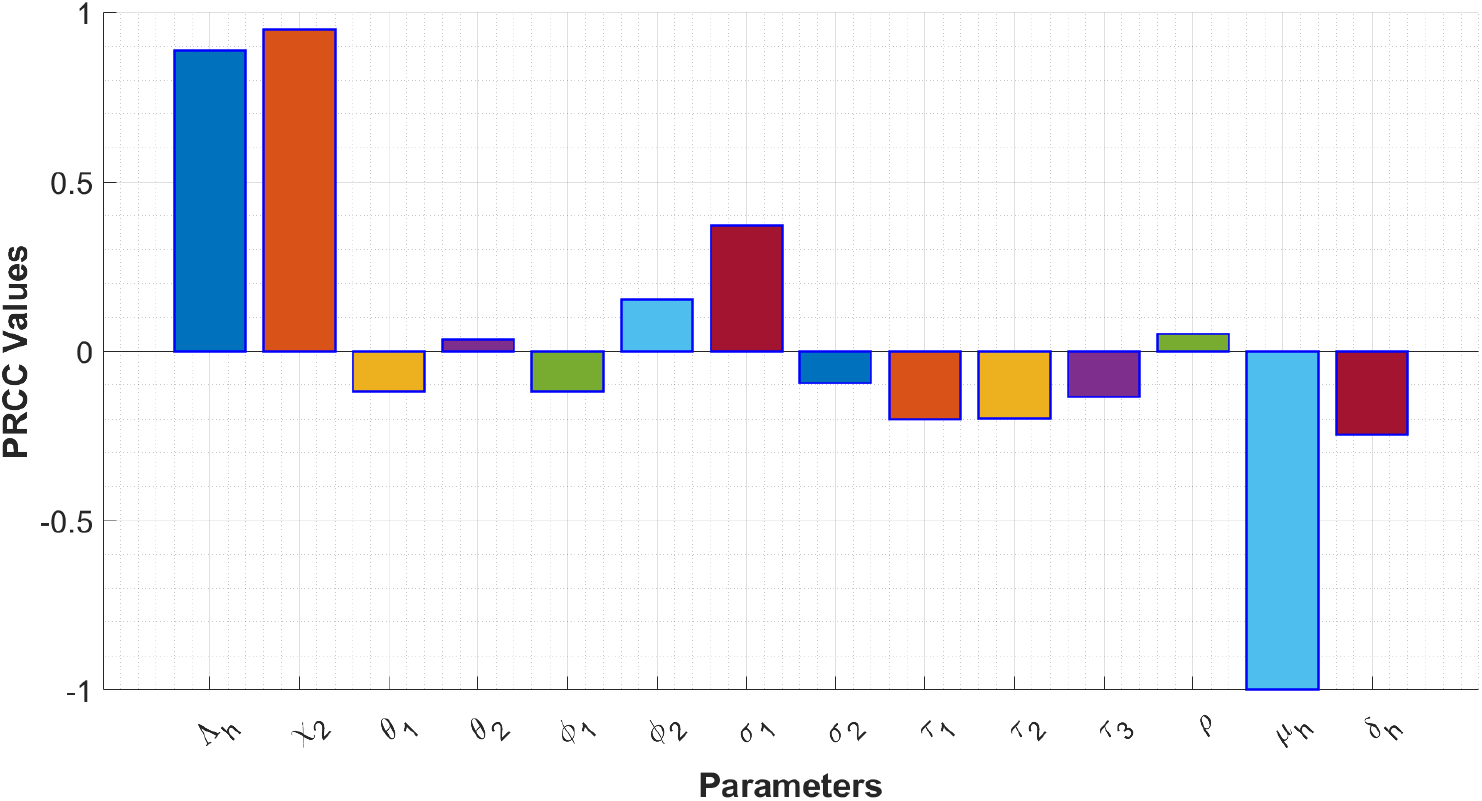
PRCC showing the impact model parameters on (*R*_*h*_).

**Figure 3:**
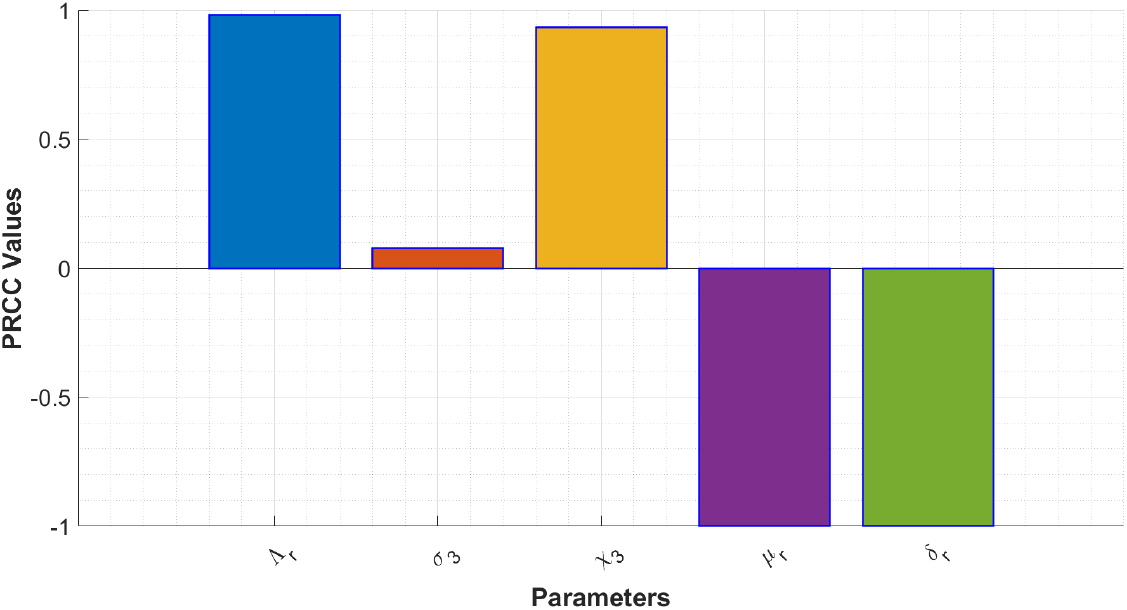
PRCC showing the impact model parameters on (*R*_*r*_).

## 6 Optimal control problem and its analysis

To devise strategies for suppressing the disease into nonexistence, we utilize the principles of optimal control theory. For evaluating the prerequisites essential to establish the attainable optimal control within the Mpox framework (1), Pontryagin’s maximum principle [47] is implemented. It is observed that vaccination of the susceptible demographic, isolation of exposed individuals, deployment of highly effective vaccines, and administration of smallpox treatments to infected persons have been strongly advocated during the Mpox epidemic. Consequently, to incorporate additional control strategies into the analysis, we modify the model (1) by diminishing the transmission coefficient by a variable factor *u*. Moreover, we introduce vaccination control denoted as *u*_*v*_ for the susceptible cohort, a control variable *u*_*q*_ for quarantining individuals, an enhancement in vaccine potency represented by *u*_*e*_, and a treatment control denoted as *u*_*t*_ for infected cases. Consequently, the newly reformulated optimal control model for the system described by (1) is as follows;

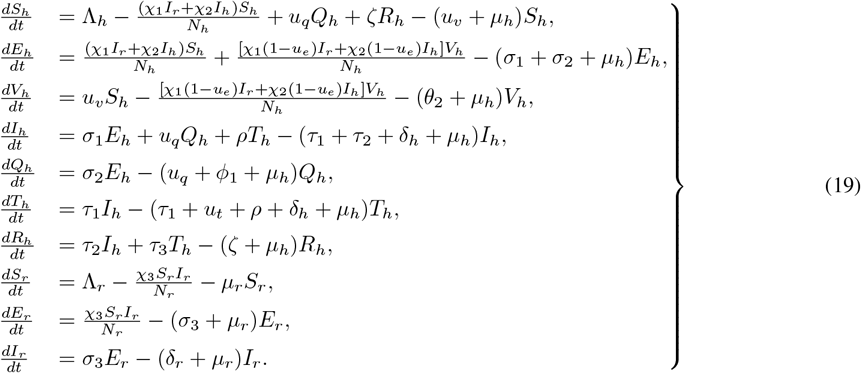

The control variables *u*(*t*) = *u*_*i*_ ∈ 𝒰, where *i* = *u*_*v*_, *u*_*q*_, *u*_*e*_, *u*_*t*_, are and are Lebesgue-measurable. The primary aim is to minimize *J* (that is, to reduce both the prevalence of infections and the expenditure associated with implementing control measures), as expressed by

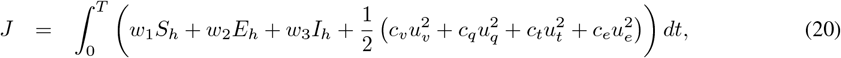

where *w*_1_, *w*_2_, *w*_3_ represent weights for human and rodent infections while *c*_*v*_, *c*_*q*_, *c*_*t*_, *c*_*e*_ represent cost coefficients for vaccination, quarantine, treatment, and vaccine efficacy improvement. We identify an optimal control set 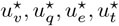such that

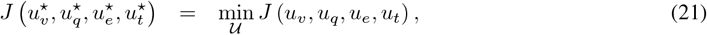

where adheres to the constraints of the model system (19). To achieve this, we employ the Pontryagin maximum principle [47], the most extensively utilized approach in optimal control modeling, to derive the solution and the requisite conditions for the optimal control problem (19).

### 6.1 Existence of optimal controls

In alignment with the findings of Fleming and Rishel as presented in [48], the existence of an optimal control quadruple minimizing (21) subject to (19) is established.

#### Theorem 5.

An optimal control *u*^⋆^ = (*u*_*v*_, *u*_*q*_, *u*_*e*_, *u*_*t*_) exists that minimizes the objective functional (20). The existence of this optimal control is demonstrated by ensuring the following conditions are satisfied:

(H1) The set of permissible controls is both convex and closed.

(H2) The dynamics of the system (19) are bounded by a linear function in terms of the state and control variables. (H3) The integrand within the objective functional (20) exhibits convexity with respect to the control variables. (H4) There exist constants *k*_1_, *k*_2_, *k*_3_ ≥ 0, and *k*_4_ ≥ 1 such that the integrand in the objective functional (20) is bounded by 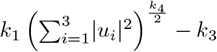

*Proof*. We construct the proof in the steps as follows:

(H1): The control set *U* = [0, 1]^4^ is a closed set. Furthermore, let *u*_*i*_, *x*∈ *U*, where *U* = (*u*_*v*_, *u*_*q*_, *u*_*e*_, *u*_*t*_) and *x* = (*x*_1_, *x*_2_, *x*_3_, *x*_4_). For any *π*∈ [0, 1], it holds that *πu*_*i*_ + (1− *π*) *x*_*i*_ ∈ *U*, thereby verifying the convexity property of the control set.

(H2): The control system (19) and its related solution are represented by Φ(*t*, Θ_1_, *u*) and Φ(*t*, Θ_2_, *u*), such that

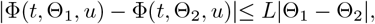

where Φ(*t*, Θ, *u*) represents the solution of the system at time *t* given the initial state Θ and the control *u*_*i*_, and *L* is a constant representing the Lipschitz continuity of the system. Also, the system of differential equations governing the transmission dynamics is

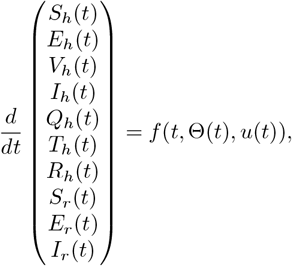

where Θ(*t*) = *S* _*h*_ (*t*) *E* _*h*_ (*t*) *V* _*h*_ (*t*) *I h* (*t*) *Q* _*h*_ (*t*) *T* _*h*_ (*t*) *R* _*h*_ (*t*) *S* _*r*_ (*t*) *E*_*r*_ (*t*) *I*_*r*_ (*t*) is the state vector, *u* _*i*_ (*t*) = *u* _*v*_ (*t*) *u* _*q*_ (*t*) *u* _*e*_ (*t*) *u* _*t*_ (*t*) ^*T*^ is the control vector (vaccination, quarantine, vaccine efficacy, and treatment). To simplify the analysis and make it more straightforward, we opt to have the kernel take the following form

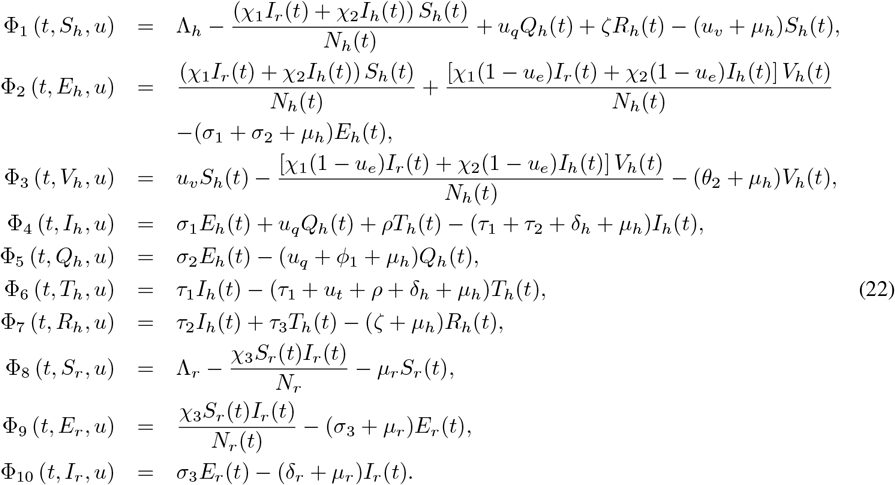

Following the above equations in (22), we obtain the constants Θ_*i*_, *i* = 1, 2, *…*, 10

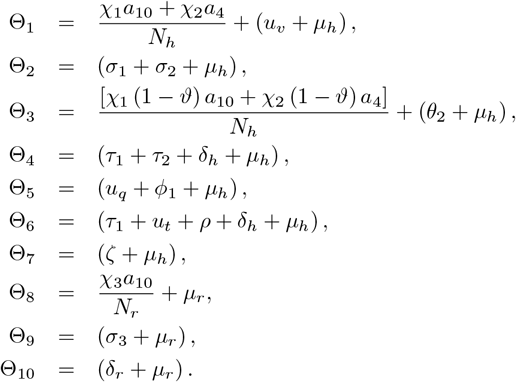

#### Theorem 6.

If assumption *H*_2_ is true and Φ_*i*_, *i* = 1, 2, *…*, 10 mollify the Lipschitz condition, then they are contractions and Θ_*i*_ ≤ 1; ∀*i* = 1, 2, *…*10.

Following Theorem (6), we first show that Φ_1_ (*t, S*_*h*_, *u*_*i*_) satisfies the Lipschitz condition. Let *S*_*h*_ and 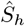 be two given functions, then

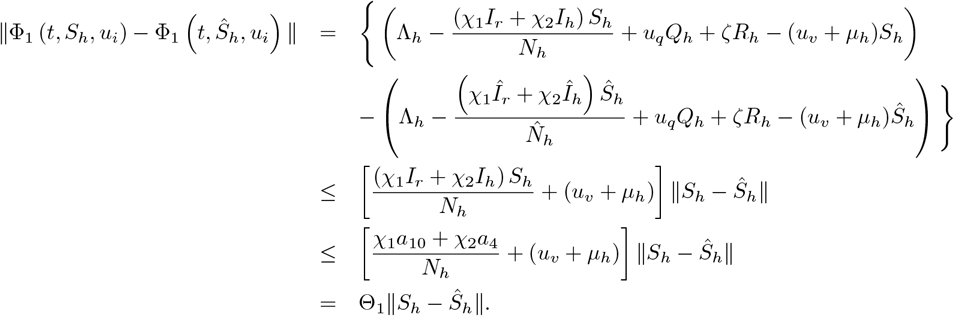

Similarly the remaining equations in (19) gives

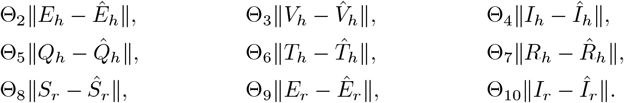

Thus, we have proven that the solution to the system is continuous with respect to the initial conditions and that the control system satisfies Condition (H2)

(H3): The objective functional for the control system is

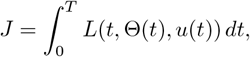

where *L*(*t*, Θ(*t*), *u*(*t*)) is the integrand, which depends on both the state vector Θ(*t*) and the control vector *u*(*t*), and *T* is the final time of the control process. We aim to show that the integrand *L*(*t*, Θ(*t*), *u*(*t*)) is convex with respect to the control variables *u*(*t*) = (*u*_*v*_(*t*), *u*_*q*_(*t*), *u*_*e*_(*t*), *u*_*t*_(*t*)). Assume the integrand has the following form

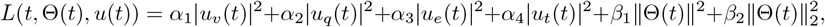

where *α*_1_, *α*_2_, *α*_3_, *α*_4_ are positive constants associated with the penalty on the control variables, *β*_1_, *β*_2_ are constants associated with the cost of the state variables, ∥Θ(*t*) ∥^2^ is a measure of the state vector Θ(*t*). This form of the integrand includes quadratic penalties on both the controls and the states. We need to prove that the integrand *L*(*t*, Θ(*t*), *u*(*t*)) is convex in *u*(*t*). Let *u*_1_(*t*) and *u*_2_(*t*) be two control inputs, and *λ* ∈ [0, 1]. We must show that

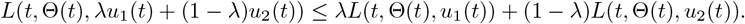

Substituting *u*(*t*) = *λu*_1_(*t*) + (1 − *λ*)*u*_2_(*t*) into the integrand

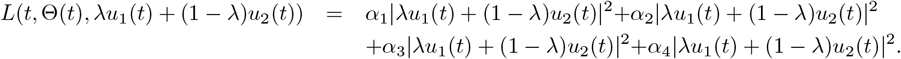

Using the property of convex functions, we know

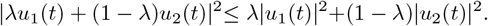

Thus, we can apply this for each term in the integrand

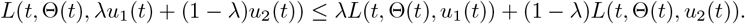

This proves that the integrand *L*(*t*, Θ(*t*), *u*(*t*)) is convex with respect to the control variables. Since *L*(*t*, Θ(*t*), *u*(*t*)) is convex in *u*(*t*), the objective functional is the integral of a convex function. Therefore, the objective functional *J* is also convex in the control variables

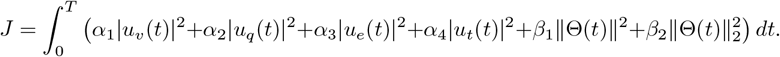

Thus, we have proven that the integrand is convex with respect to the controls, and consequently, the objective functional is convex in the control variables.

(H4): Assume the integrand has the following form

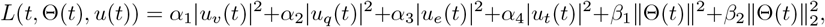

where *α*_1_, *α*_2_, *α*_3_, *α*_4_ are constants associated with the penalty on the control variables, *β*_1_, *β*_2_ are constants associated with the penalty on the state variables. To prove condition *H*4, we must show that there exist constants *k*_1_, *k*_2_, *k*_3_ and *k*_4_ ≥ 1 such that

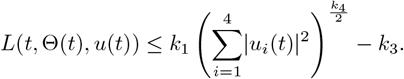

The first four terms in the integrand are quadratic in the control variables:

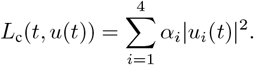

Using the inequality

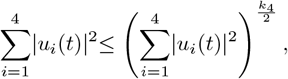

for *k*_4_ ≥ 1, we get

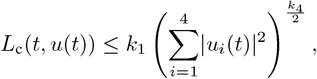

where *k*_1_ = max(*α*_1_, *α*_2_, *α*_3_, *α*_4_). The state variables Θ(*t*) contribute terms of the form

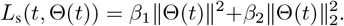

Assuming the state variables are bounded, we can find a constant *k*_2_ such that

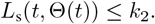

Combining the control and state terms, we get the final bound for the integrand

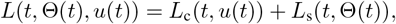

and we have

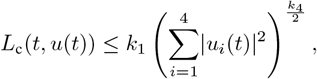

and

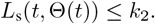

Thus, we conclude that

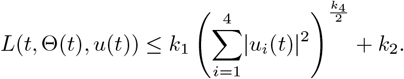

This completes the proof.

### 6.2 Characterization of Optimal Controls

Applying the Pontryagin Maximum Principle reformulates the Mpox model (19) and its corresponding objective functional (21) into the task of minimizing a Hamiltonian, *H*, with respect to the control variables *u*_*v*_, *u*_*q*_, *u*_*e*_, *u*_*t*_. The Hamiltonian for the system (19) is expressed as follows

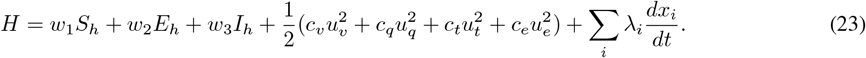

Substituting the dynamics, we expand *H* and obtain the Hamiltonian for the system to be

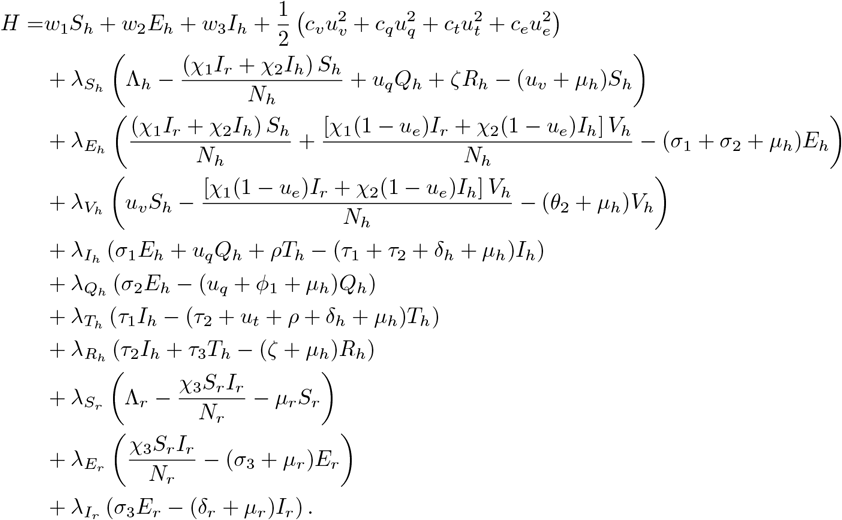

#### Theorem 7.

Let the optimal control quadruple 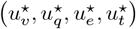 satisfy condition (21). Then, there exist adjoint variables *λ*_*i*_, corresponding to (*i* = *S*_*h*_, *V*_*h*_, *E*_*h*_, *I*_*h*_, *Q*_*h*_, *T*_*h*_, *R*_*h*_, *S*_*r*_, *E*_*r*_, *I*_*r*_), which fulfill the adjoint system described below;

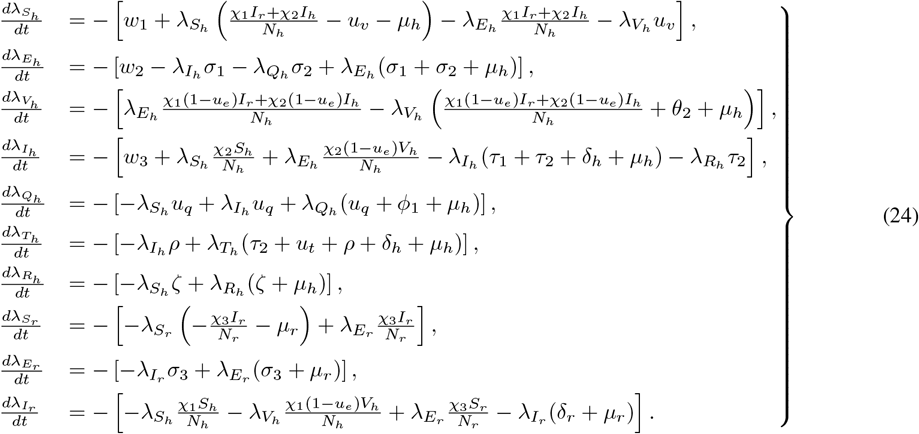

while the control functions 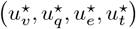satisfies the optimality condition and is given by

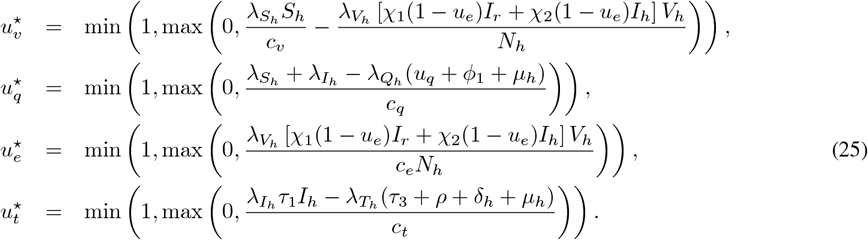

*Proof*. Referring to the Hamiltonian (23), given by

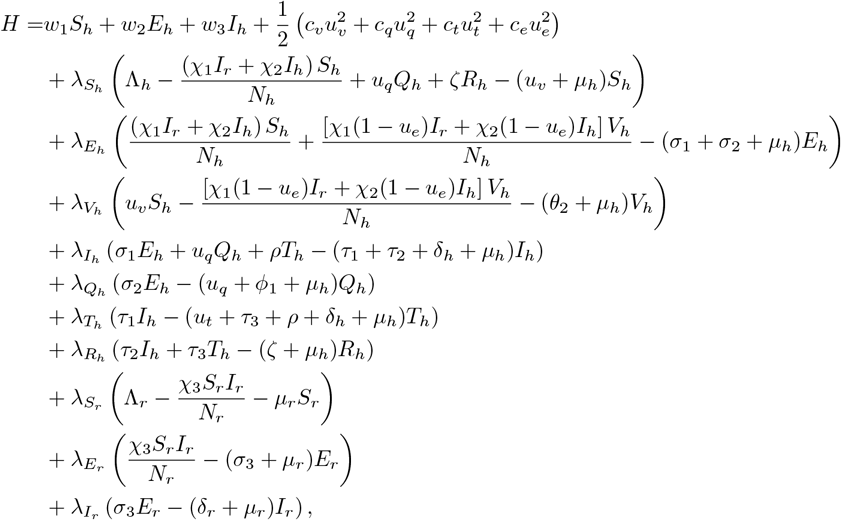

we generate the adjoint system (24) by differentiatin partially the Hamiltonain (23) with respect to the corresponding state variables *S*_*h*_, *V* _*h*_, *E* _*h*_, *I* _*h*_, *Q* _*h*_, *T* _*h*_, *R* _*h*_, *S* _*r*_, *E* _*r*_, *I* _*r*_ as 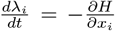. It follows that 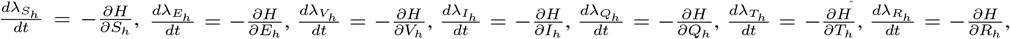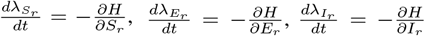. The characterization of the controls *u*^⋆^, *u*^⋆^, *u*^⋆^, *u*^⋆^ as in (21) is determined by applying the equations below;

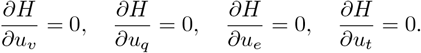

Utilizing bounds on the controls by standard argument, we determine the characterization to be in the form of (25). This completes the proof.

## 7 Numerical simulation

Model simulations are executed utilizing parameter magnitudes procured from diverse pre-existing academic references. Parameters absent in the literature are inferred based on logical proportionality postulates. An optimality framework is established by integrating the state system (19), adjoint system (24), and the optimal control characterization (25). This synthesis culminates in a two-point boundary value problem. Consequently, the optimality framework comprises a 10-dimensional system of ordinary differential equations governed by both initial and transversality conditions. The iterative scheme, advancing from forward to backward, adheres to the methodology delineated in [24], employing the fourth-order Runge–Kutta numerical method. The parameters utilized are detailed in Table 1. The investigation explores the ramifications of implementing at least two of the four optimal control variables on the dynamics of human population. The initial conditions for the state variables are specified as *S*_*h*_ = 10000, *E*_*h*_ = 2500, *V*_*h*_ = 6000, *I*_*h*_ = 1500, *Q*_*h*_ = 1000, *T*_*h*_ = 500, *R*_*h*_ = 0, *S*_*r*_ = 3000, *E*_*r*_ = 2000, and *I*_*r*_ = 1000, with the final time horizon set to 365 days. The weight coefficients of the cost functional are assigned values *w*_1_ = 200, *w*_2_ = *w*_3_ = 100, *c*_*v*_ = 1, *c*_*q*_ = 0.5, *c*_*e*_ = 0.5, and *c*_*t*_ = 1. We concluded that *w*_1_ *> w*_2_ *and w*_3_, as we assume that the cost of vaccines and implementing a vaccination program would be higher than the cost of personal quarantining exposed and treatment of infected individuals respectively. The analysis scrutinizes the influence of pairing two out of the four optimal control variables on the population dynamics of humans. Hence, we would consider model (19) parameter values in Table 1 for the simulation.

**Table 1:**
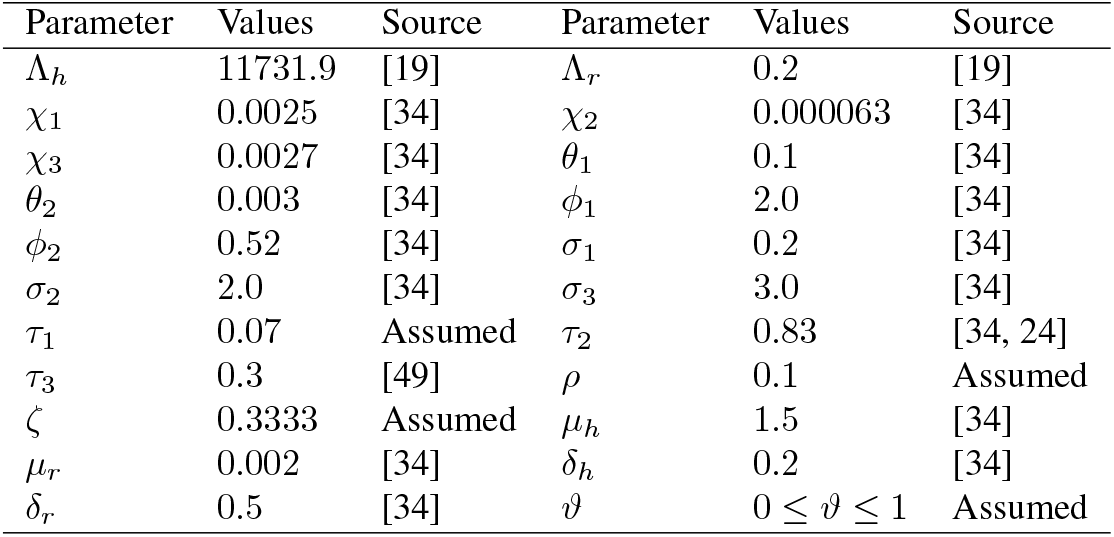
Parameters of model and their description.

| Parameter | Values | Source | Parameter | Values | Source |
| --- | --- | --- | --- | --- | --- |
| $\Lambda_h$ | 11731.9 | [19] | $\Lambda_r$ | 0.2 | [19] |
| $\chi_1$ | 0.0025 | [34] | $\chi_2$ | 0.000063 | [34] |
| $\chi_3$ | 0.0027 | [34] | $\theta_1$ | 0.1 | [34] |
| $\theta_2$ | 0.003 | [34] | $\phi_1$ | 2.0 | [34] |
| $\phi_2$ | 0.52 | [34] | $\sigma_1$ | 0.2 | [34] |
| $\sigma_2$ | 2.0 | [34] | $\sigma_3$ | 3.0 | [34] |
| $\tau_1$ | 0.07 | Assumed | $\tau_2$ | 0.83 | [34, 24] |
| $\tau_3$ | 0.3 | [49] | $\rho$ | 0.1 | Assumed |
| $\zeta$ | 0.3333 | Assumed | $\mu_h$ | 1.5 | [34] |
| $\mu_r$ | 0.002 | [34] | $\delta_h$ | 0.2 | [34] |
| $\delta_r$ | 0.5 | [34] | $\vartheta$ | $0 \leq \vartheta \leq 1$ | Assumed |

### 7.1 Optimal control simulations

In this section, we explore the ramifications of various optimal control strategies aimed at mitigating the prevalence of monkeypox within the human population. Our analysis is particularly focused on evaluating the efficacy of combined control measures in halting disease transmission. Through numerical simulations of the monkeypox model, both with and without the application of optimized interventions, we examine the impact of the control variables introduced earlier. The study investigates the effects of different combinations of optimal control strategies represented by *u*_*v*_, *u*_*q*_, *u*_*e*_, *u*_*t*_. These strategies are systematically organized into three categories: single control, double controls, and triple or quadruple controls, enabling a structured examination of the twelve potential control scenarios simulated in this research. Specifically, the strategies are defined as follows: Strategy 1 employs *u*_*v*_ and *u*_*e*_ exclusively, Strategy 2 relies solely on *u*_*q*_, Strategy 3 utilizes only *u*_*t*_, and Strategy 4 incorporates all four controls, *u*_*v*_, *u*_*q*_, *u*_*e*_, *u*_*t*_. The subsequent sections provide an in-depth simulation, analysis, and discussion of these four intervention strategies.

### 7.2 Strategy 1 (Applying controls *u*_*v*_ and *u*_*e*_ concurrently)

Figures 4 (*a*) ™ (*d*) illustrate that the application of an optimal intervention significantly reduces human vulnerability to the monkeypox virus during the intervention period compared to scenarios where adherence is absent.

**Figure 4:**
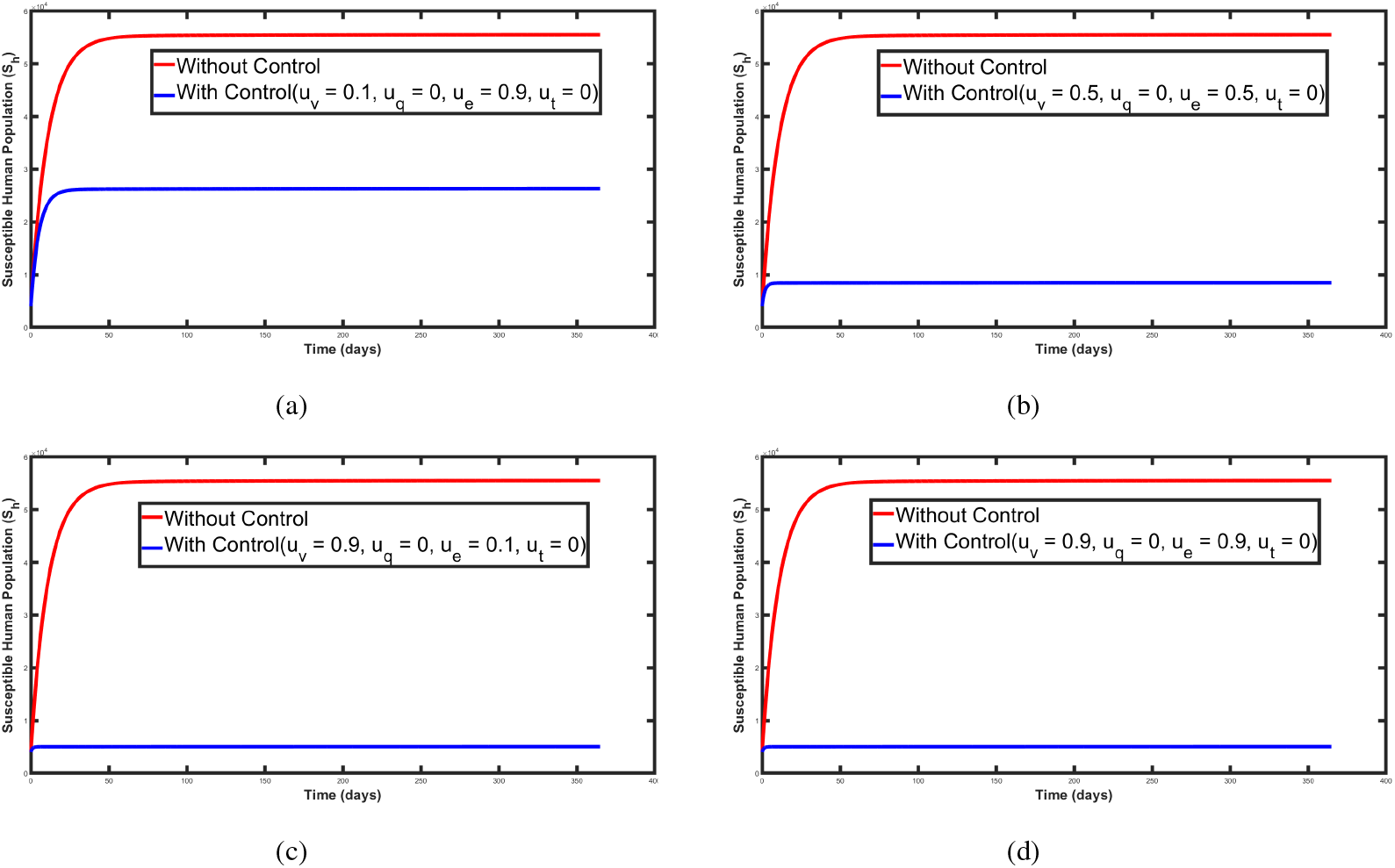
Trajectory effects of vaccination and vaccine efficacy on the susceptible human population.

Without vaccinating susceptible individuals to protect them against Mpox infection, the number of vulnerable individuals rises to a peak of 19, 677, 842.39, as shown in Figure (4*a*) and Table 2. However, introducing vaccination as a control strategy, with vaccine efficacy considered, reduces the number of susceptible individuals to 9, 474, 883. Additionally, the combined optimal deployment of *u*_*v*_ and *u*_*e*_ demonstrates maximum efficacy within a 12-day implementation period before stabilizing and asymptotically approaching zero by the end of the intervention, as depicted in Figure 4(d). This approach results in a 90.65% reduction in susceptible individuals, as detailed in Table 2.

**Table 2:** Comparison of conditions with and without control on susceptible population.

| Without control | With control | Control conditions | Percentage Change |
| --- | --- | --- | --- |
| 19677842.39 | 9474882.69 | $u_v = 0.1, u_q = 0, u_e = 0.9, u_t = 0$ | -51.87% |
| 19677842.39 | 3081401.49 | $u_v = 0.5, u_q = 0, u_e = 0.5, u_t = 0$ | -84.37% |
| 19677842.39 | 1839809.24 | $u_v = 0.9, u_q = 0, u_e = 0.1, u_t = 0$ | -90.65% |
| 19677842.39 | 1839809.24 | $u_v = 0.9, u_q = 0, u_e = 0.9, u_t = 0$ | -90.65% |

### 7.3 Strategy 2 (Executing control *u*_*q*_ independently; and synthesizing *u*_*v*_, *u*_*e*_, and *u*_*q*_ into a unified strategy)

Figure 5 elucidates that the population of exposed individuals declines more efficiently when comprehensive control measures, including immunization, vaccine efficacy, and quarantine, are employed. In the absence of adherence to these preventive protocols, the number of individuals exposed to Mpox infection is quantified at 232, 019.72. Even with an elevated quarantine rate of 90%, the exposed population remains at 232, 581.66. This underscores the ineffectiveness of utilizing quarantine as a standalone strategy for mitigating Mpox infection, as depicted in Figure 5(*a*) and detailed in Table 3. Conversely, when vaccination is amalgamated with quarantine as a combined mitigation approach, the number of exposed individuals decreases to 230, 282.83, subsequently declining further to 221, 049.79 and 213, 600.08, as illustrated in Figures 5(*b*) − 5(*d*) and elaborated in Table 3. This integration achieved a sustained maximum reduction of 7.9% in the exposed population throughout the intervention duration compared to scenarios devoid of such measures, as shown in Table 3. Moreover, it was discerned that the intervention attained peak efficacy within the initial 25 days, followed by a gradual attenuation, approaching zero effectiveness by 400 days.

**Table 3:** Comparison of conditions with and without control on exposed population.

| Without control | With control | Control conditions | Percentage Change |
| --- | --- | --- | --- |
| 232019.72 | 232581.66 | $u_v = 0, u_q = 0.9, u_e = 0, u_t = 0$ | 0.24% |
| 232019.72 | 230282.83 | $u_v = 0.1, u_q = 0.8, u_e = 0.5, u_t = 0$ | -0.75% |
| 232019.72 | 221049.79 | $u_v = 0.5, u_q = 0.5, u_e = 0.5, u_t = 0$ | -4.73% |
| 232019.72 | 213600.08 | $u_v = 0.9, u_q = 0.9, u_e = 0.9, u_t = 0$ | -7.96% |

**Figure 5:**
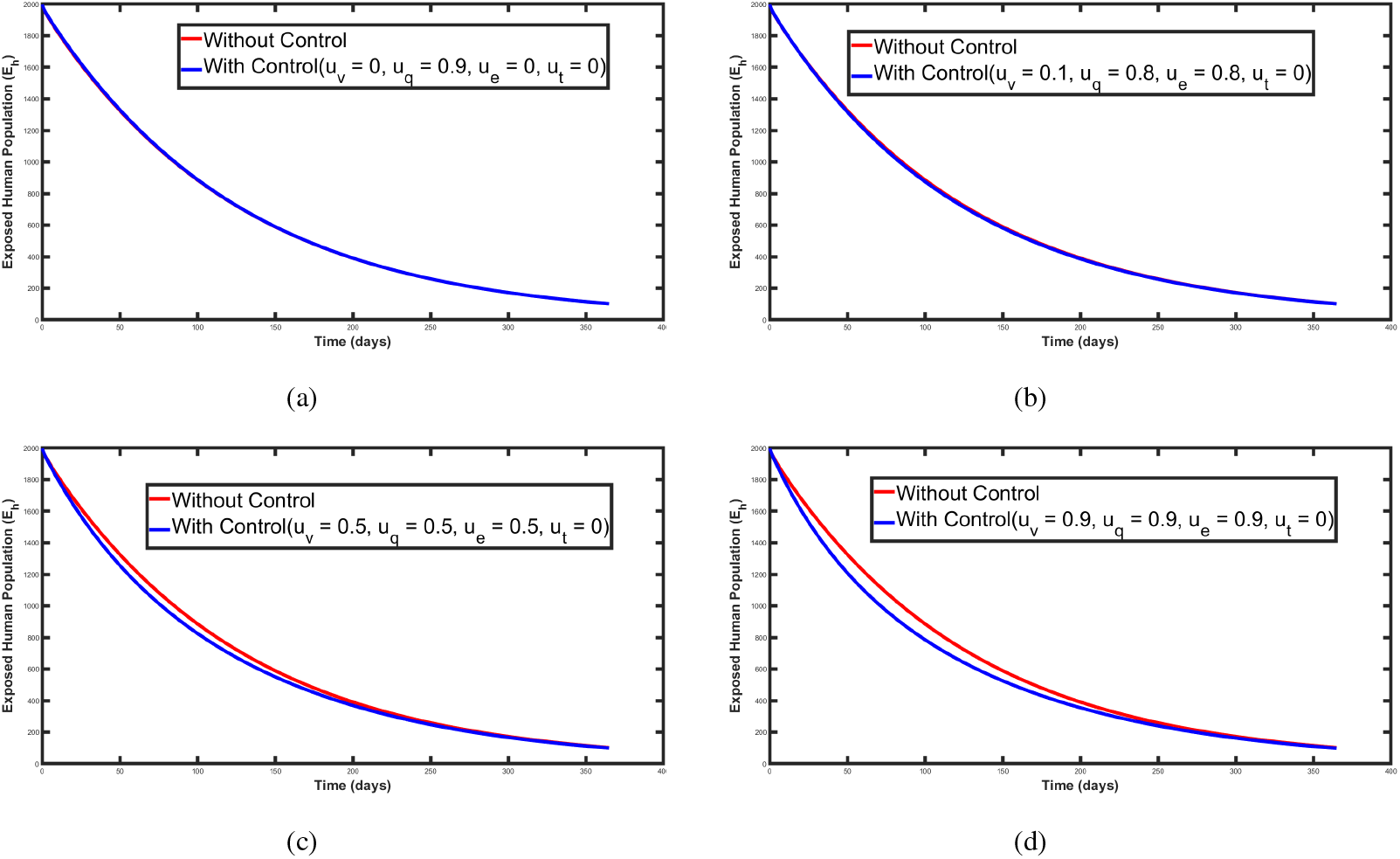
Trajectory effects of vaccination, vaccine efficacy and quarantine on the exposed human population.

### 7.4 Strategy 3 (Enforcing controls *u*_*t*_ in isolation; integrating *u*_*q*_ and *u*_*t*_; and amalgamating *u*_*v*_, *u*_*e*_, *u*_*q*_, and *u*_*t*_)

Figure 6 elucidates that the prevalence of infected individuals diminishes markedly when an amalgamation of control measures, including immunization, vaccine efficacy, quarantine, and therapeutic interventions, is sustained. In Figure 6(*a*), it becomes evident that the isolated implementation of treatment as a mitigation strategy is inconsequential to the dynamics of Mpox transmission, as the infected population continues to ascend from 810, 719.17 to 810, 951.57, despite the utilization of therapeutic measures to mitigate the contagion. Conversely, the concomitant application of quarantine and treatment as synergistic control measures precipitates a 2.73% decline in the infected spopulation, reducing the numbers from 810, 719.17 to 788, 548.94, as delineated in Figure 6(*b*) and cataloged in Table 4. Furthermore, the incorporation of vaccination, utilizing a vaccine with elevated efficacy, alongside treatment and quarantine effectuates a substantial reduction in infected individuals to 713, 616.37, equating to an 11.97% diminution over the intervention period, as illustrated in Figures 6(*c*) ™ 6(*d*) and enumerated in Table 4. Moreover, it was observed that the intervention attained maximal efficacy within the initial 50 days, subsequently attenuating to insignificance by 365 days. This analysis underscores the inadequacy of treatment as a singular control strategy in curbing the proliferation of the monkeypox virus, as the trajectory of the model employing only therapeutic intervention closely mirrors the model devoid of any preventive measures, as portrayed in Figure 6(a).

**Table 4:** Comparison of conditions with and without control on infected population.

| Without control | With control | Control conditions | Percentage Change |
| --- | --- | --- | --- |
| 810719.17 | 810951.57 | $u_v = 0, u_q = 0, u_e = 0, u_t = 0.9$ | 0.03% |
| 810719.17 | 788548.94 | $u_v = 0, u_q = 0.9, u_e = 0, u_t = 0.9$ | -2.73% |
| 810719.17 | 713616.37 | $u_v = 0.5, u_q = 0.9, u_e = 0.5, u_t = 0.9$ | -11.97% |
| 810719.17 | 713616.37 | $u_v = 0.9, u_q = 0.9, u_e = 0.9, u_t = 0.9$ | -11.97% |

**Figure 6:**
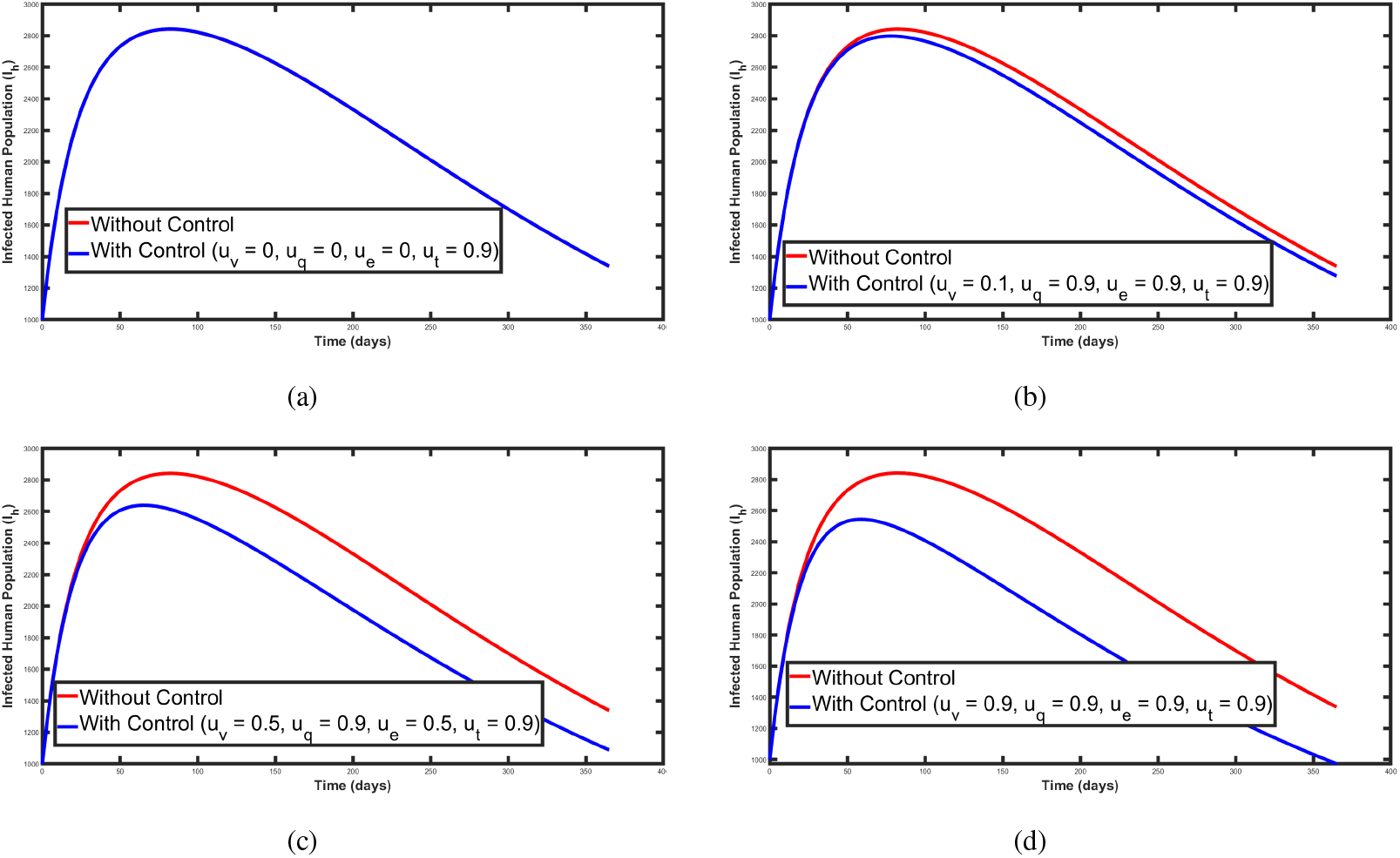
Trajectory effects of vaccination, vaccine efficacy quarantine and treatment on the infected human population.

### 7.5 Cost-effectiveness analysis

The objective in this section is to evaluate the most cost-effective strategy among the mentioned monkeypox control techniques in a setting with limited resources. To identify the most cost-effective strategy, we compare strategies 1, 2, 3, and 4. The approach outlined in related previous studies [29, 50, 51] is adopted to examine the average cost-effectiveness ratio (ACER) and incremental costeffectiveness ratio (ICER) of strategies 1, 2, 3, and 4. The ICER is the ratio of the differences in cost of two control strategies to the differences in infection averted by executing that strategy. On the other hand, the ACER is the ratio of the total cost involved in executing the strategy to the total infected prevented. The mathematical formulation

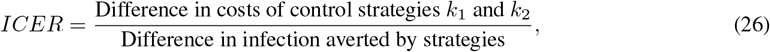

is used to exhibit the number of infections averted, and the related cost of executing the strategies under consideration in increasing order of magnitude of the infection averted is presented in Table 5. From the analysis of the strategies, those with negative Total Infection Averted were excluded as they increase infections and are not optimal. Among the remaining strategies, Strategy 2 demonstrates the best cost-efficiency, particularly the combination *u*_*v*_ = 0.5, *u*_*q*_ = 0.5, *u*_*e*_ = 0.5, *u*_*t*_ = 0, which achieves 36.51 infections averted with the lowest ACER (0.05) and ICER (10.00). For higher effectiveness, Strategy 3, with *u*_*v*_ = 0.9, *u*_*q*_ = 0.9, *u*_*e*_ = 0.9, *u*_*t*_ = 0.9, achieves the greatest reduction in infections (144,978.53) but at a significantly higher cost, reflected by a high ACER (100.10) and ICER (100.70). A balanced choice is found in Strategy 2, with *u*_*v*_ = 0.9, *u*_*q*_ = 0.9, *u*_*e*_ = 0.9, *u*_*t*_ = 0, achieving 61.30 infections averted with good cost-efficiency (ACER = 0.09, ICER = 10.72). Therefore, the optimal strategy is Strategy 2, *u*_*v*_ = 0.9, *u*_*q*_ = 0.9, *u*_*e*_ = 0.9, *u*_*t*_ = 0, offering a favorable trade-off between significant infection reduction and cost-effectiveness.

**Table 5:** Comparison of Strategies with Total Infection Averted, Total Cost, ACER, and ICER.

| Strategies | Total Infection Averted | Total Cost | ACER | ICER | Control conditions |
| --- | --- | --- | --- | --- | --- |
| **Strategy 1** | 2.33 | 5350.18 | 1.18 | -148.41 | $u_v = 0.9, u_q = 0, u_e = 0.1, u_t = 0$ |
| | 0.25 | 5205.10 | 1.15 | -803.03 | $u_v = 0.1, u_q = 0, u_e = 0.9, u_t = 0$ |
| | 1.31 | 5277.64 | 1.16 | -209.02 | $u_v = 0.5, u_q = 0, u_e = 0.5, u_t = 0$ |
| | 2.33 | 5496.18 | 1.21 | -211.08 | $u_v = 0.9, u_q = 0, u_e = 0.9, u_t = 0$ |
| **Strategy 2** | -1.87 | 164.29 | 0.02 | -87.73 | $u_v = 0, u_q = 0.9, u_e = 0, u_t = 0$ |
| | 36.51 | 365.10 | 0.05 | 10.00 | $u_v = 0.5, u_q = 0.5, u_e = 0.5, u_t = 0$ |
| | 5.75 | 365.10 | 0.05 | 63.54 | $u_v = 0.1, u_q = 0.9, u_e = 0.9, u_t = 0$ |
| | 61.30 | 657.18 | 0.09 | 10.72 | $u_v = 0.9, u_q = 0.9, u_e = 0.9, u_t = 0$ |
| **Strategy 3** | -232.40 | 81260565.33 | 100.20 | 101.42 | $u_v = 0, u_q = 0, u_e = 0, u_t = 0.9$ |
| | -540.59 | 81189631.72 | 100.08 | -87.62 | $u_v = 0, u_q = 0.9, u_e = 0, u_t = 0.9$ |
| | 97102.80 | 71425393.29 | 100.09 | 101.04 | $u_v = 0.5, u_q = 0.9, u_e = 0.9, u_t = 0$ |
| | 144978.53 | 66637847.86 | 100.10 | 100.70 | $u_v = 0.9, u_q = 0.9, u_e = 0.9, u_t = 0.9$ |

### 7.6 Discussion of results

A pioneering mathematical framework was formulated and rigorously analyzed in this study to elucidate the mechanisms governing monkeypox transmission dynamics. The model is structured upon a system of ordinary differential equations (ODEs), compartmentalized into ten distinct categories and stratified across two demographic subgroups. The human population comprises compartments for susceptible individuals, vaccinated individuals, exposed individuals, infected individuals, quarantined individuals, treated individuals, and recovered individuals.

Additionally, the rodent population is subdivided into three distinct categories: susceptible rodents, exposed rodents, and infected rodents. Unlike pre-existing models in scholarly literature, this framework incorporates the impact of imperfect vaccination, which arises from the use of vaccines originally designed to combat smallpox.

An analytical exploration of the model yielded both the disease-free equilibrium and a unique endemic equilibrium state. The next-generation matrix approach was employed to calculate the control reproduction number (*R*_*c*_), while the linearized Jacobian method established the local asymptotic stability of the model. Furthermore, the Castillo-Chavez methodology was leveraged to ascertain the global asymptotic stability of the disease-free equilibrium state, while the Lyapunov direct method was used to determine the global asymptotic stability of the endemic equilibrium. The analysis identified critical parameters influencing monkeypox dynamics, including human-to-human transmission rates, rodent-to-rodent transmission rates, progression rates, and vaccination rates. A non-autonomous version of the model incorporating optimal control strategies was derived using Pontryagin’s Maximum Principle. The influence of the optimal control set, encompassing a vaccination intervention (*u*_*v*_), quarantine measures (*u*_*q*_), enhanced vaccine efficacy (*u*_*e*_), and treatment interventions (*u*_*t*_), was scrutinized. The findings revealed that time-dependent interventions significantly curtailed monkeypox propagation across the population.

The qualitative findings were further validated through a numerical analysis using the forward-backward sweeps method, embedded within the fourth-order Runge-Kutta algorithm, to generate solutions for the optimal control problem. The simulation results demonstrated the efficacy of various control strategies in mitigating the spread of monkeypox among susceptible, exposed, and infected cohorts. Strategy 1, which focused on the dual controls of vaccination and vaccine efficacy, exhibited a pronounced impact on reducing the number of susceptible individuals, as evidenced by retrospective analyses of the disease’s trajectory. Strategy 2, incorporating controls for vaccination, quarantine, and vaccine efficacy, yielded results consistent with those of Strategy 1 for exposed cohorts. Conversely, Strategy 3, which combined vaccination, quarantine, vaccine efficacy, and treatment controls, significantly suppressed infection levels. Graphical representations revealed a considerable decline in the infected population under Strategy 3, particularly in scenarios where vaccination played a pivotal role. However, simulations without vaccination control demonstrated an early surge in exposed and infected individuals, followed by gradual suppression in the long term. The results underscore that vaccines serve as an indispensable pharmaceutical intervention but are constrained by challenges such as viral mutations and the waning efficacy of vaccines over time, compounded by limited vaccination capacity. This emphasizes the need for multifaceted intervention protocols, as reinfections following vaccination and treatment have been documented in multiple studies. Policymakers are thus urged to adopt a pragmatic approach when relaxing other mitigating measures for monkeypox.

Finally, a cost-effectiveness analysis was conducted to evaluate the economic viability of the proposed strategies. The analysis revealed that Strategy 2 exhibited the highest cost-efficiency, particularly with parameter values of *u*_*v*_ = 0.5, *u*_*q*_ = 0.5, *u*_*e*_ = 0.5, and *u*_*t*_ = 0, achieving 36.51 infections averted at the lowest Average Cost-sEffectiveness Ratio *ACER* = 0.05 and Incremental Cost-Effectiveness Ratio *ICER* = 10.00. For scenarios prioritizing maximal effectiveness, Strategy 3 (*u*_*v*_ = 0.9, *u*_*q*_ = 0.9, *u*_*e*_ = 0.9, *u*_*t*_ = 0.9) achieved the greatest reduction in infections 144, 978.53 but incurred a substantially higher cost *ACER* = 100.10; *ICER* = 100.70. A balanced approach was observed with Strategy 2 (*u*_*v*_ = 0.9, *u*_*q*_ = 0.9, *u*_*e*_ = 0.9, *u*_*t*_ = 0), which averted 61.30 infections while maintaining favorable cost-efficiency *ACER* = 0.09; *ICER* = 10.72. Consequently, Strategy 2 emerges as the optimal choice, offering a compelling trade-off between infection reduction and economic sustainability.

Based on these insights, this study advocates for public health authorities and stakeholders to enhance vaccination campaigns with high-efficacy vaccines, implement quarantine measures per WHO recommendations, and promote community awareness regarding the benefits of vaccination. Vaccination strategies with robust efficacy have been demonstrated to effectively curtail monkeypox infections, though they must be complemented by additional interventions to mitigate the disease comprehensively.

This investigation incorporates parameters derived from prior research, alongside several hypothesized values, implying that their application within this model may not guarantee absolute validity. Parameters for which precise values are unavailable, such as the vaccination rate (*θ*_1_), vaccine efficacy rate (ϑ), rodent-to-human transmission rate (*χ*_1_), human-to-human transmission rate (*χ*_2_), and rodent-to-rodent transmission rate (*χ*_3_), necessitate approximation. Future research endeavors should prioritize utilizing empirical data to ascertain these parameter estimates with greater accuracy.

## 8 Conclusion

This study introduces a novel mathematical framework to understand monkeypox transmission dynamics, incorporating imperfect vaccination and immunity loss. Key findings highlight the critical role of high-efficacy vaccines, quarantine measures, and treatment in mitigating the spread of the disease. Strategy 2, combining vaccination and quarantine, emerges as the most cost-effective intervention. While the model provides valuable insights, the use of assumed parameter values emphasizes the need for empirical data in future research. Strengthened vaccination campaigns and multifaceted interventions are recommended to effectively control monkeypox outbreaks and support public health strategies.

## Data Availability

All data produced in the present work are contained in the manuscript

